# A Spanish grammaticality judgment task for neurodegenerative diseases: inflectional, transitivity, and word order comprehension in PPA and Alzheimer’s Disease

**DOI:** 10.64898/2026.08.25.26360651

**Authors:** S Mancini, N Biondo, M Calabria, C Martin, E. García-Hernández, J. Filella-Mercè, J Selma-González, J Garcia-Castro, S. Sara Rubio-Guerra, I Sala, MB Sanchez-Saudinos, SM Grasso, I. Illán-Gala, Alexandre Bejanin, A. Lleó, J. Fortea, M. Santos-Santos

**Author notes:** **Corresponding authors**: Dr. Simona Mancini Paseo Mikeletegi 69, 2^nd^ floor 20009, Donostia-San Sebastián, Spain Dr. Miguel Santos Santos Institut de Recerca Sant Pau Carrer Sant Quintí, 77-79 08041 Barcelona, Spain.

## Abstract

Impairment in the comprehension of morphosyntactic and transitivity information does not feature in current diagnostic guidelines for primary progressive aphasia (PPA) or Alzheimer’s Diseases (AD), despite research reporting delayed sensitivity or insensitivity of these clinical populations to these linguistic domains. Moreover, studies rarely compare all three PPA variants and AD within a single design, and the literature is heavily weighted toward English, whose reduced morphology may not capture the full range of comprehension difficulties these populations experience. We developed a computer-based acceptability judgment task covering comprehension of the nominal and verbal inflection paradigm in Spanish, transitivity and word order. We recruited Spanish-speaking patients diagnosed with non-fluent/agrammatic, logopenic and semantic variants of PPA and typical AD. Psychometric evaluation confirmed good sensitivity, internal consistency and moderate correlation of task accuracy with language and neuropsychological measures. The four clinical groups retained the ability to endorse grammatical sentences but showed selective difficulty rejecting unacceptable ones. AD and the three PPA variants showed impaired comprehension of inflectional and transitivity information, whereas sensitivity to word order was comparatively preserved. Exploratory analyses revealed that short-term memory, working memory, and verbal semantics were differentially associated with sentence evaluation performance within and across groups. VBM analyses identified the left posterior temporal cortex as the main neuroanatomical correlate of grammaticality judgment performance. These findings extend prior English-language research to Spanish, demonstrating that morphosyntactic and transitivity deficits are a robust and cross-linguistically consistent feature of neurodegenerative language decline, and highlighting the importance of developing language-sensitive assessment tools for underrepresented linguistic populations.

## 1. Introduction

Extracting the propositional meaning of a sentence relies on a series of mechanisms that span across several linguistic domains: from analyzing the structuring and order of constituents and decomposing their inflectional information, i.e. morphosyntax; to determining whether a verb selects direct or indirect objects, i.e. transitivity. These abilities can become significantly compromised in individuals experiencing progressive language deterioration, such as those with primary progressive aphasia (PPA) and Alzheimer’s Disease (AD). To date, research on the deterioration of morphosyntactic and verb argument selectional properties, including the development of diagnostic criteria, has focused primarily on English-speaking patients, with an emphasis on language production. Comparatively little work has examined comprehension or has targeted Romance languages with rich inflectional systems such as Spanish (see Auclair-Ouellet, 2015; van Boxtel and Lawyer, 2021; Varlokosta et al., 2024 for reviews). The present study aimed to address this gap by designing a short grammaticality judgment task and evaluating its validity for detecting morphosyntactic^1^ and transitivity deficits in Spanish-speaking individuals diagnosed with PPA and AD.

### 1.1. Morphosyntactic and transitivity processing in PPA

PPA is an acquired neurodegenerative disease that primarily affects the network of brain regions supporting language functions, while relatively sparing cortical regions that support other cognitive domains, at least at early stages of the disease (Gorno-Tempini et al., 2008, 2011a; Gorno-Tempini et al., 2004). Current criteria for diagnosis (Gorno-Tempini et al., 2011a) identify three main PPA variants, each of which presents with salient language features and anatomical correlates. The diagnosis of the non-fluent/agrammatic variant of PPA (nfavPPA, henceforth) is typically characterized by effortful, halting speech, articulatory errors (apraxia of speech), which can co-occur with expressive agrammatism, i.e. short, syntactically simple sentences, often with omission of function words and verb inflection. Neuroanatomically, nfavPPA is characterized by predominant left fronto-insular atrophy. In contrast, anomia and single-word comprehension impairment (mostly with low-frequency items), in association with bilateral (left>right) anterior temporal lobe atrophy, are considered landmarks of the *semantic variant of PPA* (svPPA). Similarly to the svPPA, a core feature of the *logopenic variant* of PPA (lvPPA) is impaired word retrieval, which affects naming and disrupts spontaneous speech, amid relatively preserved grammatical processing (Ramanan et al. 2022). However, the presence of phonological errors and the sparing of single-word comprehension critically differentiate lvPPA from svPPA. Anatomically, atrophy, hypometabolism and hypoperfusion in left temporo-parietal, including supramarginal and angular gyri, are evident in lvPPA.

Impairment in morphosyntactic and transitivity processing is not among the ‘core’ diagnostic criteria outlined in current PPA consensus criteria (Gorno-Tempini et al., 2011a), whose assessment primarily relies on sentence-picture matching, yes/no question answering, and order-following tasks to assess comprehension deficits. While appropriate for detecting impairment in the comprehension of different degrees of syntactic complexity, from active to passive and subject/object relative clauses with (ir-)reversible thematic roles, these tasks are of limited utility for assessing participants’ ability to comprehend inflectional information and access a verb’s argument information, regardless of structural complexity. In this respect, tasks that require participants to monitor the well-formedness of sentences are better suited: by manipulating phrase/sentence structure, inflectional, and transitivity information, and thus forcing the system to process errors, it is possible to assess how sensitive the comprehension system is to these cues. This is especially relevant in the assessment of speakers of richly inflected languages like Spanish, in which inflection carries several types of information, from numerosity (singular, plural), to discourse roles (speaker, addressee), tense (past, present, future) and grammatical/biological gender, which cannot be always explicitly tested in e.g., sentence-picture matching or order-following tasks. As reviewed below, a substantial body of research has documented that all three PPA variants show either delayed detection or insensitivity to morphosyntactic and transitivity information during comprehension, suggesting the importance of assessing these linguistic domains in clinical settings. However, to the best of our knowledge, no study has so far contrasted these three levels of analysis in the same experimental design and across the three PPA variants. We note that while the focus of our study is on transitivity, i.e. the number of slots selected by a verb (one, two or three, depending on whether the verb is intransitive, transitive or ditransitive), several studies reviewed below manipulate thematic information, i.e. the type of thematic roles selected by the verb. Both transitivity and thematic information are part of a verb’s selectional properties and accessing either type of information requires processing the verb in relation to its argument(s). We therefore reviewed evidence from both types of studies.

#### Non-fluent/agrammatic PPA

Using a word-monitoring task in English, Grossman and colleagues (Grossman et al., 2005) showed delayed online inflectional processing in nfvaPPA patients compared to control participants. While in the nfvaPPA group the effect of a number agreement violation persisted well after the presentation of an incorrectly inflected verb, the effect was circumscribed to the word immediately after the anomalous target in the control group. Other word-monitoring studies in English (Peelle et al., 2007) reported nfvaPPA insensitivity to morphological (e.g., the book is being \**pick*/picked by a large group of students) and word-class anomalies (The friend is being *\*reunion*/met near the big statue). In contrast, the same paradigm provided mixed evidence concerning nfvaPPA sensitivity to thematic and transitivity anomalies. Peelle et al. (2007) reported nfvaPPA patients to be able to detect thematic anomalies like “The president is being \**spiced* in the peaceful country”, while Price and Grossman (2005) reported insensitivity to both thematic and transitivity violations (“The hamster brings beside chewed carrots chips”), regardless of whether the to-be monitored word was presented immediately or some words after the incongruence. More recently, online electrophysiological data by Barbieri et al. (2021) showed that individuals with nfvaPPA are impaired in the processing of number inflection and thematic violations, as evidenced by their lower accuracy and failure to elicit a P600 effect compared to the control group. Notably, inflectional and thematic information contained in verbs has been found to be problematic for nfvaPPA also in production, which has been taken to suggest a general and modality-independent impairment in the representation of verbs’ morphosyntactic and argument structure information in this variant (Thompson & Mack, 2014; Wilson et al. 2014)

Existing investigations adopting metalinguistic tasks like grammaticality judgments did not cover the analysis of phrase structure well-formedness such as word order anomalies (but see Peelle et al. 2007 for word-class anomalies). In this respect, evidence from studies administering the Northwestern Anagram Test (NAT, Weintraub et al., 2009) can provide indirect evidence concerning the ability of patients diagnosed with nfvaPPA to process constituents and sentence structure. In the NAT, participants are presented with a picture and are required to produce a target sentence that describes it by ordering scrambled single-word cards, a process that crucially relies on metalinguistic knowledge of syntactic rules similar to what is required in a grammaticality judgment. Studies in English and other languages have shown that patients diagnosed with nfvaPPA are significantly impaired in this task (Italian: Canu et al., 2019; Japanese: Watanabe et al., 2018; English: Weintraub et al., 2009). Such impairment in the ability to monitor the structural well-formedness of a sentence could emerge also in comprehension.

In sum, evidence from English and few other languages (Italian, Japanese) suggests that individuals diagnosed with nfvaPPA can show deficits in the comprehension of a wide spectrum of linguistic domains: from inflectional to argument and phrase/sentence structure well-formedness. To the best of our knowledge, no study so far has investigated the comprehension of these three domains in Spanish.

#### Logopenic variant of PPA

Although not affected by omission of function words and telegraphic speech production, typically referred to as agrammatic output in PPA literature, individuals with lvPPA can present with significant morphosyntactic comprehension impairment, usually accompanied by paragrammatism in production (Amici et al., 2007; Gorno-Tempini et al., 2008; Gorno-Tempini et al., 2004; see review in Wilson et al., 2012). The predominantly non-agrammatic nature of lvPPA’s expressive impairment is also supported by the relatively spared performance of this PPA variant in the NAT, both in English (Weintraub et al. 2009; Thompson et al. 2012) and Italian (Canu et al. 2019). Two studies investigating morphosyntactic processing with electrophysiological paradigms have shown that logopenic patients have reduced sensitivity to inflectional violations, as shown by their attenuated P600 effects (Barbieri et al., 2021; Kielar et al., 2012). Importantly, it has been suggested that morphosyntactic impairment in the logopenic variant is mediated by short-term memory impairment (Gorno-Tempini et al. 2008). Similar to nfvaPPA, no study so far has investigated morphosyntactic and transitivity processing in Spanish speakers diagnosed with lvPPA, nor has the nature of any such impairment been tested.

#### Semantic variant of PPA

Morphosyntactic comprehension has been generally found to be spared in the semantic variant of PPA. Using a grammaticality judgment task, Cotelli and colleagues (Cotelli et al., 2007) showed that Italian individuals affected by this PPA variant are not impaired in the offline detection of anomalies involving either number agreement or sentence structure (e.g. wh-question, clitic placement). Along similar lines, Grossman et al. (2005) reported individuals affected by semantic dementia to be as sensitive as cognitively unimpaired individuals in the detection of number agreement violations in an online word-monitoring task. These findings contrast with those from an online word-monitoring task by Price and Grossman (2005), who showed that a small sample of individuals with semantic dementia was not sensitive to the presence of transitivity violations, either in the time interval immediately following the violation or several syllables down the stream.

A potential explanation for the heterogeneous results documented in English and Italian could be the different stage at which deficits were tested in these languages^2^. As the disease progresses, paragrammatism can emerge (Wilson et al., 2010, 2012), along with morphosyntactic comprehension deficits. Indeed, findings from the longitudinal follow-up of patient AK by Rochon et al. (2004) showed that over a 5-year period, her ability to evaluate the grammaticality of conceptually plural but morphologically singular sentences (e.g. “the gang of the boys fights”) was eventually compromised.

Evidence for inflectional impairment in the semantic variant of PPA also emerges from paradigms that tested processing at the word level. Using a gender decision task in Spanish, Lambon Ralph et al.(2011) tested access to gender information contained in nouns and showed that svPPA patients’ accurate performance was influenced by the regularity of the ending-to-gender association in Spanish (-o: masculine; -a: feminine). Along similar lines, the choice of the correct irregular past tense options in English was found to be impaired in individuals with svPPA (Patterson et al., 2001, Wilson et al., 2014), suggesting that impairment in the comprehension of inflectional information in svPPA may be concomitant with the presence of irregular morphology.

No study to date has investigated inflectional morphology, transitivity, or phrase/sentence structure wellformedness in svPPA, nor has the potential contribution of the underlying semantic deficit to the processing of these linguistic domains been examined.

### 1.2. Morphosyntactic and transitivity processing in AD

The majority of patients diagnosed with lvPPA have underlying AD pathology, suggesting that this PPA variant is an atypical presentation of AD (Chare et al., 2014; Leyton et al., 2017, 2017; Mesulam et al., 2014; Spinelli et al., 2017). Neuroanatomically, typical AD presents with bilateral atrophy of posterior parietal regions and medial temporal lobe structures (McKhann et al., 2011a). Clinical manifestations predominantly feature non-language centered symptoms, such as episodic memory, semantic memory, working memory and executive function impairment.

In language, similarly to lvPPA, patients with AD present with semantic impairment, which manifests itself with anomia (Leyton et al., 2017; Reilly et al., 2011) and single-word comprehension deficits. Sentence-level semantic processing also appears to be impaired, as evidenced by studies examining the processing of verb argument properties in English. These studies have reported insensitivity to thematic information (Grossman et al., 1995, 1996, 1997; Price & Grossman, 2005) but preserved sensitivity to transitivity violations (Kim & Thompson, 2004; Price & Grossman, 2005), a pattern that may be explained by AD patients’ impaired semantic processing abilities, given that thematic anomalies carry a predominantly semantic component, as opposed to the more grammatical nature of transitivity violations.

Similar to lvPPA, AD patients’ comprehension of constituent structure and order in metalinguistic tasks is understudied. To our knowledge, the only study testing this level of linguistic analysis is Cotelli et al (2007), who report preserved sensitivity of AD patients in the analysis of anomalies involving clitic placement but not of wh-questions.

A heterogeneous scenario emerges from studies adopting metalinguistic judgment tasks focusing on the analysis of inflectional information. Impaired processing of number, person tense and aspect inflection was found in Greek AD (Fyndanis et al., 2013), with errors showing participants’ preference towards accepting ungrammatical sentences as correct, a pattern also found in post-stroke aphasia (see Hagiwara, 1995; Varlokosta et al. 2006; Wilson and Saygin, 2004). Using a word-monitoring task, Grossman and Rhree (2001) reported significantly delayed sensitivity of AD patients to number agreement anomalies in English compared to the control group. However, preserved abilities to comprehend gender, person and tense inflection was found in Hebrew AD patients. Rather than reflecting cross-linguistic differences, these contrasting results can plausibly be attributed to the varying sensitivity of the different tasks and their dependent variables used across studies (i.e., accuracy in an offline judgment task, and response latencies in online word-monitoring and online reading tasks, see Varlokosta et al. 2024 for a discussion on online vs. offline task sensitivity to detect morphosyntactic impairment in AD).

As with lvPPA, the existence of a genuine morphosyntactic impairment in AD patients has been frequently called into question (see Boxtel and Lawyer, 2021; Varlokosta et al. 2024 for reviews), with some researchers proposing that poor comprehension of inflectional morphology stems from impaired working memory rather than from the progressive erosion of linguistic abilities *per se*. Using a speeded grammaticality judgment task with varying sentence lengths, Waters and Caplan (1997) found that AD patients and healthy participants were equally sensitive to the violation of anaphoric coreference. However, the AD group was slower to respond when sentences contained two noun phrases (e.g., *The woman with the man washed herself/*himself; The woman in the room washed herself/*himself*) compared to simpler sentences (e.g., *The woman washed herself/*himself*). The authors concluded that AD does not impair morphosyntactic processing *per se*, as offline paradigms might suggest, but rather working memory, resulting in difficulties with longer sentences regardless of their syntactic complexity (see also Waters et al., 1991).

To our knowledge, no existing studies in Spanish have yet tested AD’s sensitivity to inflectional, structural and transitivity well-formedness in the same experimental task.

## 2. The current study

Impairment in the comprehension of morphosyntactic and transitivity information is not described in PPA and AD classification guidelines. Nevertheless, as reviewed above, a consistent body of studies has reported either delayed sensitivity or insensitivity in the comprehension of these linguistic domains across the three variants of PPA and in AD. This suggests that deficits in these domains could instead represent a common diagnostic trait in language- and non-language centered neurodegenerative diseases, perhaps due to different underlying non-linguistic deficits. The lack of studies comparing the three PPA variants and AD within the same experimental design, together with the prevalence of data from English, an inflectionally poor language, makes it difficult to identify the full scope of the comprehension impairment in these neurological conditions, the underlying neuroanatomical correlates, and potential variant-specific behavioral differences. The current study sought to fill this gap by testing and contrasting the ability to comprehend morphosyntactic and transitivity information of four groups of Spanish speakers diagnosed with PPA or AD. Specifically, we assessed the diagnostic utility of a short acceptability judgment task that covers a wide range of inflectional information expressed in Spanish nouns and verbs - person, number, gender and tense agreement-along with within-constituent word order and transitivity manipulations. Word order, inflectional and transitivity manipulations allowed us to test fundamental facets of sentence comprehension while keeping the focus on syntactic properties of nouns and verbs, thus avoiding manipulations that would trigger a semantic analysis of the sentence, as happens with thematic role anomalies. Finally, with VBM analyses we aimed to identify patterns of atrophy that were most consistently associated with performance in our task.

From a psychometric perspective, we expected our behavioral task to have enough sensitivity and specificity to discriminate the three PPA variants and AD groups from the cognitively healthy group, as well as to show converging validity with tasks traditionally used to assess general sentence comprehension and production (e.g., sentence-picture matching, sentence elicitation, sentence repetition) and divergent validity from neuropsychological tasks measuring unrelated cognitive domains (e.g., visuo-perceptual/spatial abilities).

To evaluate the extent of deficits and identify potential group-specific patterns, we compared the accuracy and response times of the three PPAs and AD groups in grammatical and ungrammatical conditions with those of the cognitively healthy group. We expected all three PPA variants and the AD group to show sizeable deficits in detecting ungrammaticality, given converging evidence from prior studies in English and other languages documenting insensitivity to morphosyntactic and transitivity manipulations in both PPA (Barbieri et al. 2021; Grossman et al. 2005; Kielar et al. 2018; Peele et al. 2007; Price and Grossman, 2005; Wilson et al. 2012; see Lambon-Ralph et al. 2011 for a gender-decision task in Spanish) and AD (Fyndanis et al. 2013). In addition, we expected accuracy to be more affected in ungrammatical compared to grammatical conditions, as suggested by studies with neurodegenerative and post-stroke aphasia that reported predominant impairment in ungrammatical sentence evaluation, as opposed to within-normal-limits performance on grammatical stimuli (Fyndanis et al. 2013; Hagiwara, 1995; Wilson and Saygin, 2004).

Findings from neuroimaging studies with cognitively healthy Spanish speakers also provide supporting evidence for our hypothesis that sizeable impairments in ungrammaticality detection could be present across our four clinical groups. The analysis of morphosyntactic anomalies (relative to correct sentences) in Spanish has been found to rely on a cortical network that involves left inferior (pars orbitalis, triangularis and opercularis) and middle frontal areas, temporal (posterior and anterior middle temporal gyrus) and inferior parietal (including the angular gyrus) regions (Carreiras et al., 2015; Mancini et al., 2017; Quiñones et al., 2014, 2018). Atrophy of these brain regions could therefore lead to poorer accuracy in our clinical groups (compared to the cognitively healthy group).

While we expected the processing of ungrammaticality to be affected both in language-dominant and language non-dominant neurodegeneration, general cognitive functions such as short-term, working memory and verbal semantics could have a different effect across groups given their differences in cognitive impairment profiles and brain atrophy patterns. In this respect, auditory short-term and working memory deficits should account for impairment in lvPPA and AD, respectively, while generally degraded conceptual processing could be associated with poor ungrammaticality comprehension in svPPA and AD. Working memory could also be a significant predictor of ungrammaticality processing in the nfvaPPA group, given the involvement of frontal areas in this cognitive function and in nfavPPA brain atrophy (see review in Eikelboom et al. 2018).

Finally, we tested whether any diagnostic group was particularly impaired in detecting a specific type of grammatical violation. Insensitivity to number, tense and transitivity manipulations was expected in the three PPA variants, based on studies in English (Barbieri et al. 2021; Grossman et al. 2005; Peele et al. 2007; but see Cotelli et al. 2007 for Italian, who found no number agreement impairment in svPPA). Existing evidence on the processing of gender and person agreement is less consistent across the literature, largely due to the scarcity of studies examining PPA in morphologically rich languages like Spanish. Nevertheless, we predicted that impaired sensitivity to gender and person agreement might emerge in the logopenic and semantic variants of PPA, in line with fMRI studies conducted in Spanish reporting left posterior/inferior parietal and anterior temporal activation during the processing of these features (Mancini et al. 2017; Quiñones et al. 2014). Inferior parietal activation during sentence processing has been interpreted as reflecting the retrieval of multiple types of linguistic information, including phonological, morphological, and lexico-syntactic information (Hagoort, 2013; Hagoort and Indefrey, 2014; see discussion in Quiñones et al. 2018), while anterior temporal lobe engagement during morphosyntactic processing has been linked to the mechanisms that bind inflectional and semantic-conceptual information, as is the case for person and gender (e.g., first-person inflection signals the presence of a speaker in the sentence; the suffix -a marks feminine biological gender in Spanish; see discussion in Mancini et al. 2017; Quiñones et al. 2017). As for AD, similar to PPA, and in keeping with previous studies (Fyndanis et al. 2013; Grossman and Rhee, 2001), we expected generalized impairment across number, person and tense, which could be ascribed to the synergistic effect of working memory limitations and degraded semantic-conceptual representations of inflectional information (Fyndanis et al., 2013; Waters & Caplan, 1997).

Available grammaticality judgment studies did not include phrase structure anomalies in their materials. We hypothesized that impairment in phrase structure would emerge most prominently in patients diagnosed with nfvaPPA, based on evidence from the NAT (Thompson et al. 2012; Weintraub et al. 2009; see Canu et al. 2019 for its adaptation in Italian), in line with models attributing left inferior frontal areas a key role in basic syntactic structure operations like combining two elements to form a new constituent during comprehension (Friederici, 2017). The lvPPA group could also show insensitivity to within-constituent word order manipulations. Recently, Matchin and Hickok (2019) have associated posterior temporal regions to expressive paragrammatism and deficits in syntactic structure building mechanisms that emerge in terms of altered word order.

## 3. Methods

### 3.1. Participants

We recruited 72 participants from the Sant Pau Initiative on Neurodegeneration (SPIN cohort) that were prospectively evaluated at the Sant Pau Memory Unit (Barcelona, Spain) between 2021 and 2024. We included patients with a diagnosis of one of the three variants Primary Progressive Aphasia (pwPPA N=39, logopenic variant [lvPPA]=16, semantic [svPPA]=10, non-fluent/agrammatic [nfavPPA]=13), Alzheimer’s disease dementia (AD, N=11), and cognitively normal controls (CN) who completed the experimental acceptability judgment task (N=22). Patients were diagnosed based on internationally accepted criteria (Gorno-Tempini et al., 2011b; McKhann et al., 2011b), had normal or corrected-to-normal vision and no hearing impairment. Control participants had normal cognitive scores in a standard neuropsychological evaluation and a Clinical Dementia Rating scale (Morris, 1994) sum of boxes score of 0. All participants received a neurological and neuropsychological evaluation, provided CSF and plasma samples for biomarker analysis, and underwent neuroimaging (MRI and/or PET FDG and/or CT scan) which was reviewed by a behavioral neurology specialist for establishment of clinical diagnosis. Specific details on the SPIN cohort evaluation protocol are described elsewhere (Alcolea et al., 2019).

All participants gave written consent, and the ethics committee of Hospital Sant Pau approved all procedures included in this study, in accordance with the Declaration of Helsinki. The demographic and linguistic profile of each group is illustrated in Table 1. Participants in the five groups were speakers of Spanish who had acquired this language either as a native language, before or simultaneously with Catalan.

**Table 1.** Demographic and linguistic profile of the five groups. Significant differences relative to the CN groups are indicated in bold.

|  | CN<br>(N=22) | AD<br>(N=11) | lvPPA<br>(N=16) | nfavPPA<br>(N=13) | svPPA<br>(N=10) | p.value |
| --- | --- | --- | --- | --- | --- | --- |
| **Gender (%)** |  |  |  |  |  | .26 |
| Female | 9 (40.9%) | 7 (63.6%) | 5 (31.2%) | 6 (46.2%) | 7 (70%) |  |
| Male | 13 (59.1%) | 4 (36.4%) | 11 (68.8%) | 7 (53.8%) | 3 (30%) |  |
| **Age**<br>**(median, Q1, Q3)** | 67.8 (62.2, 73.4) | 70.8 (66.7, 76.1) | 75.417 (67.3, 78.3) | 75.8 (69, 78.2) | 73.3 (72.5, 76.8) | .06 |
| **Handedness (%)** |  |  |  |  |  | .26 |
| Ambidextrous | 0 (0%) | 1 (9.1%) | 3 (18.8%) | 0 (0%) | 1 (10%) |  |
| Left | 0 (0%) | 1 (9.1%) | 1 (6.2%) | 0 (0%) | 0 (0%) |  |
| Right | 21 (100%) | 9 (81.8%) | 12 (75.0%) | 13 (100%) | 9 (90%) |  |
| **Years of education**<br>**(median, Q1, Q3)** | 20 (16.2, 20) | **11 (9.5, 17.5)\*** | 16 (11.8, 20)* | **12 (11, 14)\*** | **11.5(9, 15)\*** | **.005** |
| **Years post onset**<br>**(median, Q1, Q3)** |  | 5 (3.1, 7) | 3.1 (2.5, 4.2) | 3.2 (1.9, 3.7) | 6.8 (3.7, 7.4) | **.03\*** |
| **Language profile**<br>**(%)** |  |  |  |  |  | .22 |
| Balanced bilingual | 3 (13.6%) | 7 (63.6%) | 3 (18.8%) | 3 (23.1%) | 3 (30.0%) |  |
| Bilingual - Catalan<br>Dominant | 8 (36.4%) | 2 (18.2%) | 5 (31.2%) | 6 (46.2%) | 3 (30.0%) |  |
| Spanish<br>monolingual | 11 (50.0%) | 2 (18.2%) | 7 (43.8%) | 4 (30.8%) | 3 (30.0%) |  |
| Bilingual - Spanish<br>dominant | 0 (0.0%) | 0 (0.0%) | 1 (6.2%) | 0 (0.0%) | 1 (10.0%) |  |

Legend: CN=Cognitively normal; AD = Alzheimer's Disease; lvPPA = logopenic variant PPA; nfavPPA = non-fluent/agrammatic variant PPA; svPPA=semantic variant PPA. Balanced bilingual = Spanish and Catalan acquired simultaneously; Bilingual - Catalan/Spanish dominant = earlier acquisition and/or greater exposure to Catalan/Spanish; Spanish monolingual = participant that only speaks Spanish. P.values are from ANOVA or Kruskal-Wallis tests. \*ANOVA for years post onset only includes the four diagnostic groups

### 3.2. Cognitive and linguistic assessment

Individuals from both the PPA, AD and CN groups underwent a standard one-hour neuropsychological evaluation to assess episodic verbal memory, visual memory, attention, executive functions, visuospatial, visuoperceptual and visuoconstructive functioning and language as previously described <u>(Alcolea et al., 2019)</u>. Neuropsychiatric symptoms, functional impact, and the level of global cognitive impairment were also assessed (see Table 2).

**Table 2.** Neurocognitive assessment.

|  | **CN**<br>**(N=22)**<br>% correct<br>(Q1, Q3) | **AD**<br>**(N=11)**<br>% correct<br>(Q1, Q3) | **lvPPA**<br>**(N=16)**<br>% correct<br>(Q1, Q3) | **nfavPPA**<br>**(N=13)**<br>% correct<br>(Q1, Q3) | **svPPA**<br>**(N=10)**<br>% correct<br>(Q1, Q3) | **p.value** |
| --- | --- | --- | --- | --- | --- | --- |
| **Global cognition** |  |  |  |  |  |  |
| MMSE (/30) | 29 (28, 29.8) | 24 (22, 25)* | 24.5(21.5, 27.2)* | 27 (26, 28)* | 23.5 (21, 28.5)* | < .001 |
| **Language** |  |  |  |  |  |  |
| Boston Naming Test (/60) | 93.3 (88.3, 96.7) | 75 (66.7, 80)* | 40 (30, 60.8)* | 80 (65, 90)* | 43.3 (28.3, 65)* | <.001 |
| Synonyms (/100) | 95 (90.6, 97.5) | 85 (75, 86.2)* | 85 (75, 87.5)* | 85 (67.5, 88.8)* | 80 (75.6, 90) | <.001 |
| CERAD Order Comprehension (/15) | 15 (15, 15) | 15 (15, 15) | 14 (12.8, 15) | 15 (13, 15) | 15 (15, 15) | 0.061 |
| Sentence matching test (/12) | 100 (100, 100) | 95.8 (91.7, 100) | 91.7 (83.3, 100) | 91.7 (66.7, 100) | 100 (91.7, 100) | 0.125 |
| Sentence elicitation test (/18) | 83.3 (75, 91.7) | 58.3 (33.3, 79.2)* | 50 (44.4, 54.2)* | 38.9 (33.3, 55.6)* | 61.1 (33.3, 61.1)* | .002 |
| Category verbal fluency (# animals/60 sec.) | 29 (27, 38) | 16 (13.5, 23.2)* | 12.5 (8.8, 16.2)* | 16 (11, 19)* | 14 (6, 17)* | <.001 |
| Phonetic verbal fluency ("p" words/60 seconds) | 27 (22.2, 34.8) | 16.5 (10.2, 23.8)* | 11 (7, 16)* | 11 (5, 12)* | 13 (5, 18)* | <.001 |
| **Executive functions** |  |  |  |  |  |  |
| TMT A (time to complete in sec.) | 39.5 (33, 53.5) | 62.5 (46, 116)* | 72 (47, 128.5)* | 62 (51, 91)* | 50 (45, 63) | <.001 |
| TMT B (time to complete in sec.) | 79.5(66.5, 102.8)* | 650 (489.5, 700)* | 218 (183, 700)* | 260 (235, 334)* | 206 (95, 650)* | <.001 |
| Forward digit span | 6 (5, 6) | 5 (4.250, 5.8)* | 4 (3, 5.2)* | 4 (4, 5)* | 5 (5, 5.8) | .003 |
| Backward digit span | 4.5 (4, 5) | 3 (3, 4)* | 3 (2, 4)* | 3 (3, 4)* | 3.5 (3, 4)* | <.001 |
| **Visuo-spatial, perceptive, and constructive function** |  |  |  |  |  |  |
| Number Location VOSP (/10) | 9.5 (8.2, 10) | 5.5 (2.8, 7.8)* | 8 (6, 9)* | 9 (8, 10) | 8 (6.2, 9)* | .001 |
| Poppelreuter figures (/10) | 10 (10, 10) | 10 (9.250, 10) | 9 (7.8, 10)* | 10 (9, 10)* | 9 (8, 9.8)* | <.001 |
| Clock drawing test (/10) | 9.5 (7.6, 10) | 6 (4.2, 7.250)* | 6.5 (5, 7.9)* | 9 (7, 9.5) | 7.5 (4.4, 8.9)* | <.001 |
| CERAD Figure Copy (/11) | 11 (11, 11) | 9 (7.5, 10)* | 10 (9, 11)* | 10 (8, 11)* | 9 (6, 11)* | <.001 |
| **Visual memory** |  |  |  |  |  |  |
| CERAD Figures recall (/11) | 8.5 (7.2, 10) | 0 (0, 0.8)* | 3 (0.5, 4.8)* | 6 (2.5, 8)* | 1 (0, 2)* | <.001 |
| **Verbal memory** |  |  |  |  |  |  |
| FCSRT immediate free recall (/48) | 26.5 (21.2, 29.8) | 4 (2.2, 12)* | 5.5 (1.8, 10)* | 13 (10, 21)* | 4 (0, 7.2)* | <.001 |
| FCSRT immediate cued recall (/48) | 46 (44.2, 47) | 16.5(13.2, 22.8)* | 20 (9.5, 25.5)* | 32 (15, 39)* | 10.5 (3.2 23)* | <.001 |
| FCSRT delayed free recall (/16) | 11 (8.5, 13.8) | 0 (0, 2.2)* | 0.5 (0, 4.5)* | 5 (4, 8)* | 0.5 (0, 2.5)* | <.001 |
| FCSRT delayed cued recall (/16) | 16 (15.2, 16) | 5.5 (2.8, 7.8)* | 7 (1.8, 11)* | 11 (10, 14)* | 4.5 (0.2, 8)* | <.001 |

Legend: CERAD: Consortium to Establish a Registry for Alzheimer's Disease; FCSRT: Free and Cued Selective Reminding Test; VOSP: Visual Object Space and Perception Battery. MMSE: Mini Mental State Examination. TMT: Trail Making Test. P.values are from ANOVA or Kruskal-Wallis tests. \*Indicate significant difference between CN and each diagnostic group.

Briefly, global cognitive and functional status is assessed with the Mini Mental State Examination (MMSE, Blesa et al., 2001) and Clinical Dementia Rating (CDR) (Morris, 1994). Language and semantic memory are assessed with the 60-item Boston Naming Test (BNT), the Order Comprehension test from the Consortium to Establish a Registry for Alzheimer’s Disease (CERAD) battery, and the semantic (1 min, animals) and phonemic (1 min, ‘p’) fluency tests (Peña-Casanova et al., 2009). Executive functions are evaluated with the Digits Span Forward and Backward test from the Wechsler Memory Scale and Trail Making Test parts A and B (TMT; Tombaugh, 2004).

Visuospatial, visuoperception, and visuoconstructive abilities are assessed with the Number Location subtest from the Visual Object Spatial Perception (VOSP) battery, Poppelreuter overlapping figures test, and the Clock-Drawing (to oral command) test and CERAD figures and Complex Rey Figure copy tests, respectively. Visual memory is evaluated using the CERAD figures and Complex Rey Figure recall. Finally, verbal memory is assessed using the Free and Cued Selective Reminding Test (FCSRT; Grau-Guinea et al., 2021) and (CERAD) word list.

In addition to the standard SPIN protocol, participants underwent supplemental linguistic assessments. These included an adaptation to Spanish of the multicenter standardized SpeechFTLD protocol (Baqué et al., 2022), Grammatical comprehension and production were evaluated with an adaptation to Spanish of the UCSF sentence to picture matching task (Wilson et al., 2010) and a picture sentence elicitation task (Varkanitsa et al., 2024) respectively. Single word comprehension was evaluated with an adaptation to Spanish of a synonym task that required selection of a target word among two distractors (Noonan et al., 2010).

### 3.3. Acceptability Judgment task

The material included in the grammaticality judgment task comprised 48 sentences, half of which were ungrammatical. Grammaticality was manipulated by creating inflectional, transitivity and word order anomalies within sentence constituents, as illustrated in Table 3.

**Table 3.**
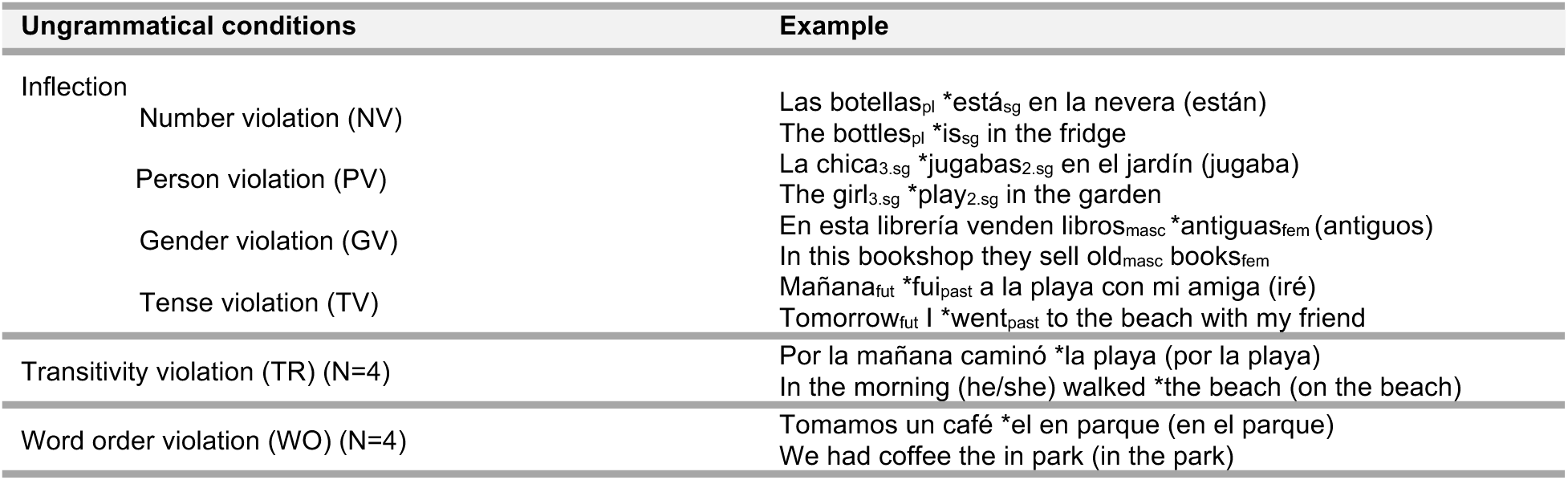
Sample of experimental material (correct version in parenthesis)

Inflectional anomalies included the manipulation of different features (number, gender, person and tense) in article-noun, noun-adjective, noun-verb and adverb-verb agreement relations. The distance between the controller (e.g. article/subject/adverb) and the target (noun/verb) of an agreement relation was manipulated to create 8 adjacent and 8 distant anomalous relations. Transitivity was manipulated in four sentences by making transitive verbs intransitives and vice versa. Finally, the order of within-phase constituents was manipulated to create word order anomalies.

### 3.4. Procedure

All patients were tested in a computer-based version of the task implemented in Psychopy (version 2021.1.4) on a Lenovo Ideapad C340 laptop. Participants were visually and auditorily presented with Spanish sentences on a computer screen and were instructed to evaluate their acceptability by pressing either one of two buttons on a keyboard (1= correct, 0=incorrect). Participants were explained that they should evaluate whether the sentence they just read/listened to contained a grammatical error or was acceptable in their Spanish, with acceptable implying that they would say it the same way in a non-experimental setting. Trial presentation was randomized across participants.

Participants were allowed to respond without any time limitation and were encouraged to take a break whenever needed, in addition to the break scheduled after 24 trials. Simultaneous auditory and visual presentation was meant to reduce reliance of task performance on a specific modality therefore reducing confounding effects of potential mild deficits in either one.

### 3.5. Neuroimaging data: acquisition and processing

#### MRI acquisition

Neuroimaging data was available for a subsample of 35 PwPPA (13 nfvaPPA, 13 lvPPA, 9 svPPA), 11 PwAD, and 22 CN. Participants were scanned using a Siemens Prisma Fit 3T scanner with the Syngo MR E11 Software and using a 32-channel head coil at IDIBAPS, Barcelona (Spain). High-resolution T1 images were acquired using a volumetric magnetization prepared rapid gradient echo (MPRAGE) sequence [repetition time (TR) = 2300 ms; echo time (TE) = 2.98 ms; inversion time (TI) = 900 ms; slice thickness = 1.0 mm; acquisition matrix = 256 x 256; voxel size = 1.0 x 1.0 x 1.0 mm].

#### MRI preprocessing

*Voxel-based morphometry*. First, raw DICOM scans were converted to the Neuroimaging Informatics Technology Initiative format using MRIcroGL software (www.nitrc.org/projects/mricrogl). T1-weighted images were then processed and analyzed with the voxel-based morphometry (VBM) pipeline implemented in the Computational Anatomy Toolbox (CAT12.9 r2560) (www.neuro.uni-jena.de/cat) for Statistical Parametric Mapping (SPM12, v7771) (www.fil.ion.ucl.ac.uk/spm/software/spm12) running on MATLAB R2022b (the MathWorks, Inc., Natick, Massachusetts, United States). The VBM pipeline consists of several stages (tissue segmentation, spatial normalization to a standard Montreal National Institute template, modulation, and smoothing), as previously described (Kurth et al., 2015). Grey matter (GM) segmented images were normalized, modulated, and smoothed with a 8-mm Gaussian kernel.

### 3.6. Statistical analysis

#### General Cognitive and Language Assessment

Normality of the general cognitive and language assessment scores was tested with the Shapiro-Wilk test. Depending on the distribution of each variable, group comparisons were performed by ANOVA followed by Tukey’s post hoc test corrected for multiple comparisons or the Kruskal-Wallis test followed by Dunn’s Test for multiple comparisons (adjusted for multiple comparisons using Benjamini-Hochberg correction). We used Chi^2^ or Fisher’s exact tests to assess differences in categorical variables. These and all following statistical analyses described were performed in R statistical software (2024.12.1). Results are reported in Table 2.

#### Acceptability judgment task

The psychometric properties of the task were evaluated in terms of its construct validity, specificity and sensitivity, and its internal consistency. Construct validity was examined by assessing the convergent and divergent validity of our task with, respectively, established tests that assess sentence comprehension and production, and tests that evaluate memory, visuo-spatial abilities and global cognition, by performing Spearman correlations.

To determine task specificity (i.e. the potential of the task to recognize participants without comprehension impairment) and sensitivity (i.e. the potential of the task to recognize participants with comprehension impairment), we generated Receiver Operating Characteristics (ROC) curves for the comparison of the PPA and the AD groups with the control group (Table 4) using the pROC function in R (Robin et al., 2011).

**Table 4.** Measures of diagnostic accuracy.

|  | AD vs. CN | nfavPPA vs. CN | lvPPA vs. CN | svPPA vs. CN |
| --- | --- | --- | --- | --- |
| AUC (95% CI) | 0.74<br>(0.54-0.98) | 0.75<br>(0.55-0.95) | 0.84<br>(0.70-0.99) | 0.85<br>(0.70-0.99) |
| Threshold | 0.93 | 0.91 | 0.8 | 0.85 |
| Specificity | 0.64 | 0.73 | 1 | 0.91 |
| Sensitivity | 0.73 | 0.77 | 0.62 | 0.7 |
| Youden's index | 0.37 | 0.5 | 0.62 | 0.61 |

*Notes.* AUC - Area Under the Curve

The reliability of our task was assessed by computing the internal consistency (Chronbach’s alpha), i.e. the correlation among the items of a test, to evaluate the extent to which they measure the same underlying construct. Both general correlation among items and among experimental conditions was measured, to assess whether the entire list of items and their grouping into different experimental conditions measured the same underlying construct.

In addition to psychometric tests, three sets of regression analyses were conducted. We first assessed performance differences in accuracy and response times among the control, the three PPA and AD samples, controlling for differences in disease severity as measured by MMSE. Accuracy and RT data from the grammaticality judgment task were analyzed with (generalized) linear mixed effect models in R, using the (g)lmer functions available in the lme4 package (Bates et al., 2015). For both dependent variables, models of increasing complexity in the fixed-effect structure were built by sequentially entering predictors of interest in a forward fashion and testing their explanatory power using likelihood ratio tests (LRTs). Random-factor structure was kept constant by including by-participants and by-items random intercepts. Numerical predictors were centered to avoid convergence and collinearity issues. Post-hoc comparisons were conducted on estimated marginal means (EMM, henceforth) and corrected with False Discovery Rate with the function emmeans (Lenth and Piaskowski, 2026). The simplest model included socio-demographic predictors, i.e. years of education and years post onset, which could not be balanced across groups. Predictors of interest - namely Group (5 levels: CN, svPPA, lvPPA, nfvaPPA and AD), Grammaticality (grammatical, ungrammatical) and the interaction between them-were sequentially entered into the model, to assess their explanatory power, followed by MMSE. Results from LRTs are illustrated in Table 5 and Figure 5. A supplementary analysis to assess sentence length effects on accuracy and response times was run with a set of regression models that included the number of words per sentence as predictors, along with group, grammaticality and MMSE (See Supplementary Materials 3).

Subsequently, following a similar statistical approach, we conducted a series of exploratory analyses aimed to identify potentially group-specific cognitive factors predicting accuracy in PPA and AD, as well as diagnosis-specific accuracy patterns. Given the small size of our groups, the outcomes of these analyses are to be regarded as hypothesis-generating, rather than conclusive. Nevertheless, they are clinically and theoretically relevant given the lack of studies investigating morphosyntactic and transitivity comprehension in Spanish-speaking individuals with neurodegenerative diseases. We opted to focus on accuracy only, as this dependent variable is the gold standard for linguistic assessment and allows for easier measurement in clinical settings. Centered scores from short-term, working memory and conceptual semantics tasks were entered as fixed-effect factors in GLMEMs, together with grammaticality, group and their interaction with each cognitive test to predict accuracy. To control for disease severity and demographic differences among the 4 clinical groups, the simplest model included MMSE scores, language dominance and gender. Fixed effects and their interactions were entered in the models sequentially, as illustrated in Table 6. Three-way interactions were disentangled by testing the effect of cognitive scores on the grammaticality effect within each group using the emtrends function (Lenth and Piaskowski, 2026) in R.

For the second exploratory analysis, condition (7 levels: correct, gender, number, person, tense, transitivity and word order violations) was included as fixed factor, along with group and their interaction. The simplest model included age and years of education. Here we report the results of the analysis of accuracy for its greater relevance compared to response times in clinical settings (for the results of the analysis of RTs, see Supplementary Materials 3)

### Neuroimaging data

#### Grey matter comparison across diagnostic groups

Whole brain analyses of differences in GM were investigated using an analysis of variance (ANOVA) test across groups, including age and total intracranial volume as nuisance variables. Statistical maps were thresholded at p < 0.001 (uncorrected) at the voxel level with a minimum cluster extent threshold of k = 50. Resulting T-maps were visualized using Surf Ice software for rendering, with colour indicating T-statistics from the regression analysis (Figure 1).

**Figure 1.**
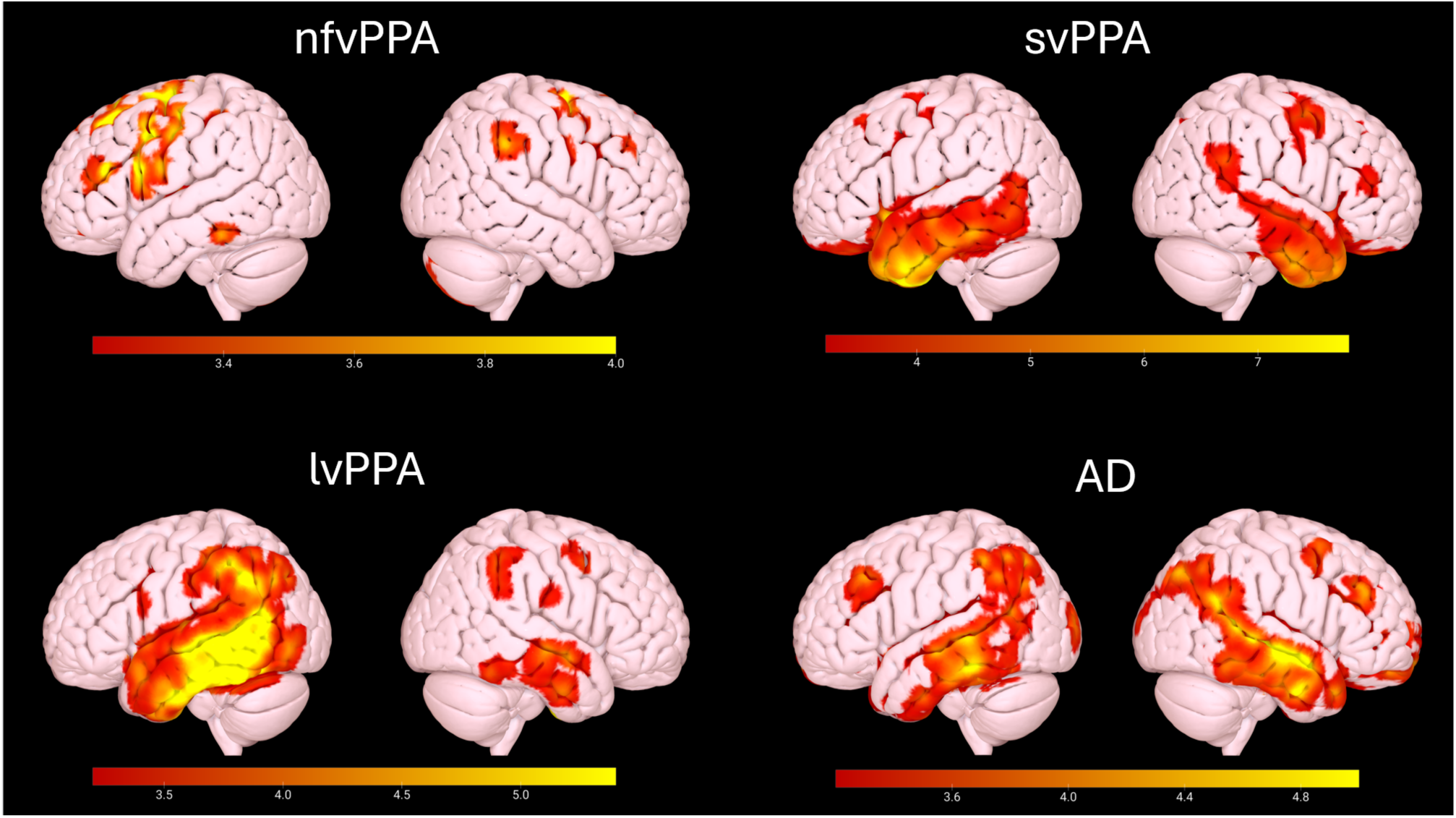
Decreased grey matter volume in all patient groups compared to the control group. Age and total intracranial volume were included as covariates and maps are thresholded at p < 0.001 uncorrected with a cluster extent threshold k=50. Colors indicate T-values, with warmer colors reflecting stronger group differences.

#### Imaging correlates of performance on the experimental task

Whole brain voxel-wise multiple linear regression in SPM12 was used to assess the relationship between performance on the experimental task and GM volume in patients. Statistical models were corrected for age and total intracranial volume. Statistical maps were thresholded at p < 0.001 (uncorrected) at the voxel level with a cluster extent threshold of k = 50. The location of significant clusters was assessed using the Neuromorphometrics atlas. Resulting T-maps were visualized using Surf Ice software for rendering, with colour indicating T-statistics from the regression analysis.

We conducted four different analyses. First, we identified the GM voxels associated with total accuracy in all patients (Figure 2, panel A). To partial out the effects of short-term auditory and working memory, we ran the same regression covarying for digits forward (Figure 2, panel B) and backward (Figure 2, panel C) span, respectively. Finally, to investigate if correlates differed according to etiology, we ran separate regressions in patients with an etiologic diagnosis of AD (lvPPA and AD, Figure 2, panel D) and FTLD (nfvPPA and svPPA, Figure 2, panel E).

**Figure 2.**
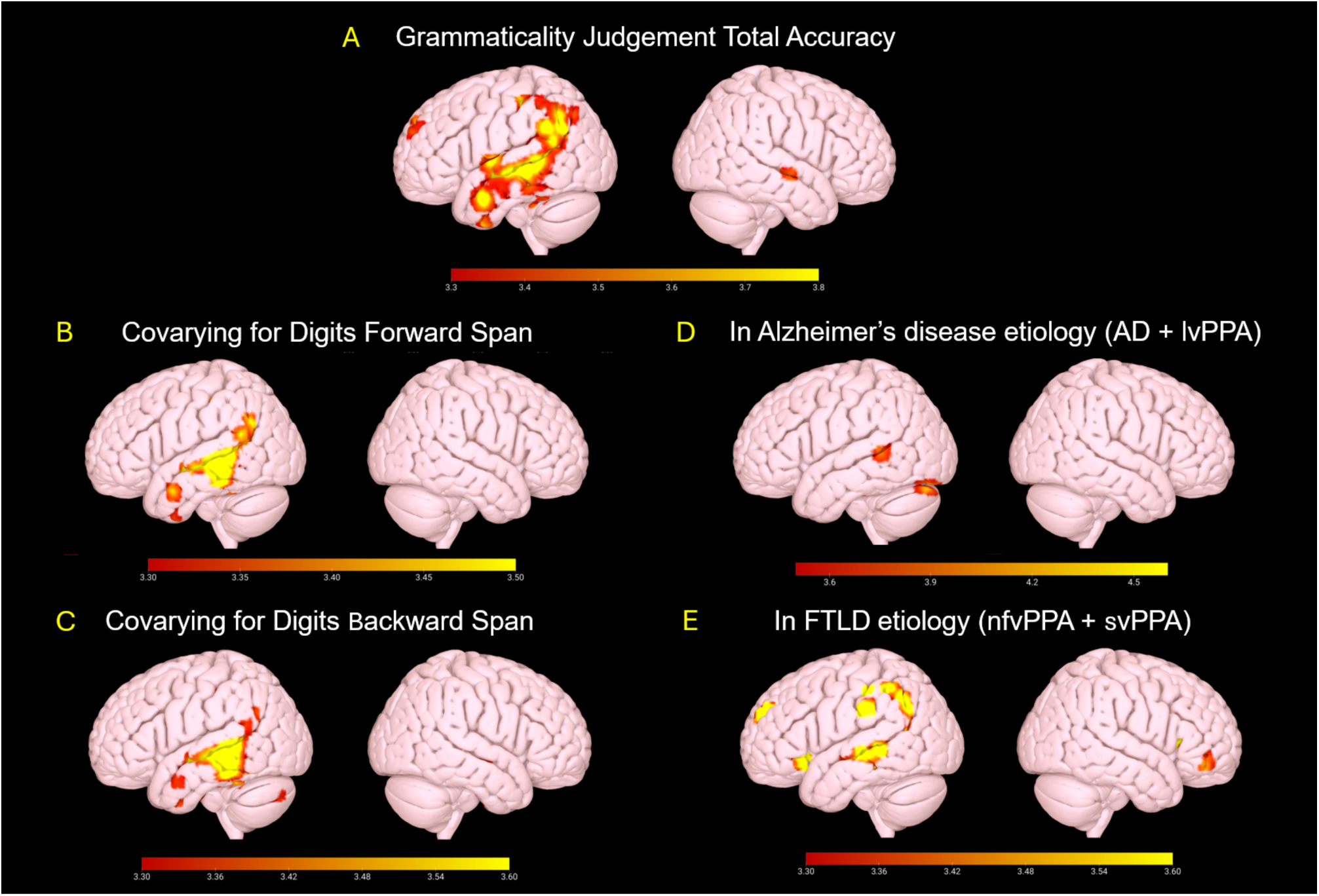
GM voxels correlated with total accuracy in patients (A), after covarying for scores on digits forward (B) and backward (C) spans. Correlates in patients with an etiologic diagnosis of Alzheimer’s Disease (AD + lvPPA) (D) and Frontal Temporal Lobar Degeneration (nfvPPA + svPPA) (E). Age and total intracranial volume were included as covariates and maps are thresholded at p < 0.001 uncorrected with a cluster extent threshold k=50. Colours indicate T-values, with warmer colors reflecting stronger group differences.

## 4. Results

### 4.1. Construct validity and reliability

#### Convergent and divergent validity

Correlations for convergent and divergent validity are illustrated in Table 3. Overall, the assessment of construct validity over the clinical sample showed a moderate positive correlation between grammaticality judgment accuracy and tests assessing sentence comprehension and elicitation. No significant correlation emerged either between grammaticality judgment accuracy and RTs or between RTs and language comprehension and elicitation.

Divergent validity analysis revealed that accurate evaluation of sentence grammaticality positively correlated with forward and backward digit span, as well as with measures of global cognition like the MMSE. In contrast, grammaticality judgment accuracy negatively correlated with TMT tests, such that better abilities to evaluate the grammaticality of a sentence were associated with faster completion of both TMT tests.

No significant correlation emerged between grammaticality judgment RTs and cognitive tests.

**Table 3.** Convergent and divergent validity of GJ accuracy and response times with other tasks measuring language and cognitive abilities. Significant correlations are indicated in bold.

|  |  | Accuracy | RTs |
| --- | --- | --- | --- |
| Convergent validity | Accuracy | 1 | -0,22 |
|  | RTs | -0.22 | 1 |
|  | Sentence matching | **0.46** | -0,06 |
|  | Sentence elicitation | **0.53** | -0,13 |
|  | CERAD Order comprehension | **0.38** | -0,28 |
| Divergent validity | TMT-A | **-0.34** | 0,19 |
|  | TMT-B | **-0.47** | 0,19 |
|  | CERAD Figure copy | **0.40** | -0,20 |
|  | CERAD Figure recall | 0.25 | -0.29 |
|  | Poppelreuter | 0.28 | -0,06 |
|  | Clock Drawing task | 0.25 | -0,12 |
|  | VOSP Number Location | 0.14 | -0,02 |
|  | MMSE | **0.35** | -0,20 |
|  | Forward Digit Span | **0.44** | -0,15 |
|  | Backward Digit Span | **0.42** | -0,19 |

Notes. RTs= response times.

#### Reliability

Chronbach’s alpha revealed excellent correlation among items (Chronbach’s alpha: .89, 95% CI: .84-.92) and deletion of any single item would not substantially improve overall reliability (range of α if item deleted: .88-.89). Similarly, correlation among conditions was excellent (Chronbach’s alpha: .84, 95% CI: .77-.89), with α ranging from .79 to .85 after single item deletion.

#### ROC analysis

Measures of diagnostic accuracy are reported in Table 4 and illustrated in Figure 3 below.

**Figure 3.**
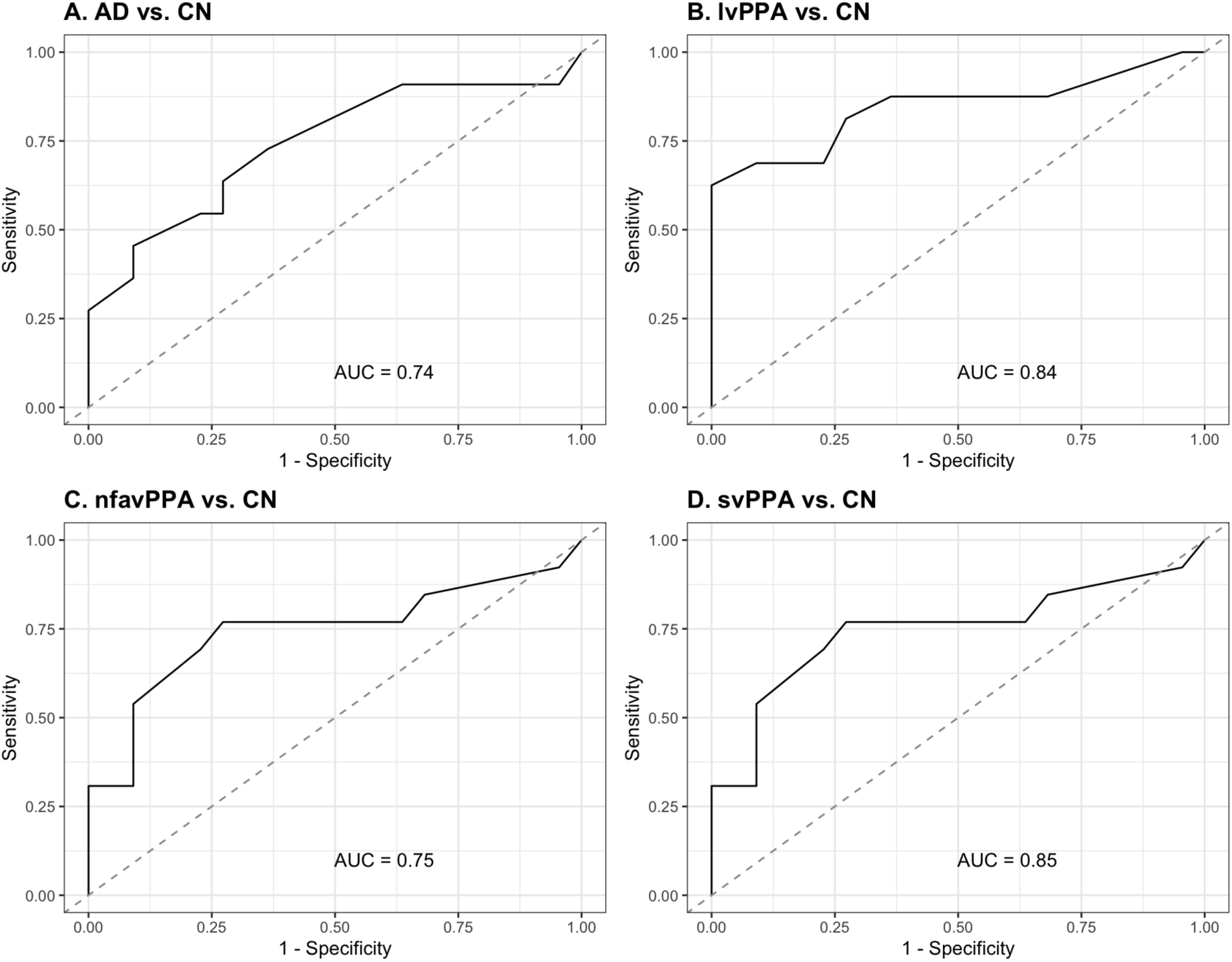
ROC curves for the discrimination between the four clinical groups and the cognitively non-impaired group. Legend: AD: Alzheimer’s Disease; nfvaPPA: nonfluent/agrammatic variant PPA; lvPPA: logopenic variant PPA; svPPA: semantic variant PPA; CN: cognitively normal.

ROC analysis yielded AUCs ranging from .74 to .85, indicating fair to good power to discriminate the CN from the four diagnostic groups. The test showed overall fair sensitivity to detect impairment in all clinical groups except for lvPPA, for which sensitivity was below 0.7. Similarly, specificity to identify cognitively non-impaired participants was fair-to-very good, apart from the comparison between AD and CN.

### 4.2. Accuracy and response times performance

Accuracy and RTs performance across groups are illustrated in Figure 4 and Table 3 below.

**Figure 4.**
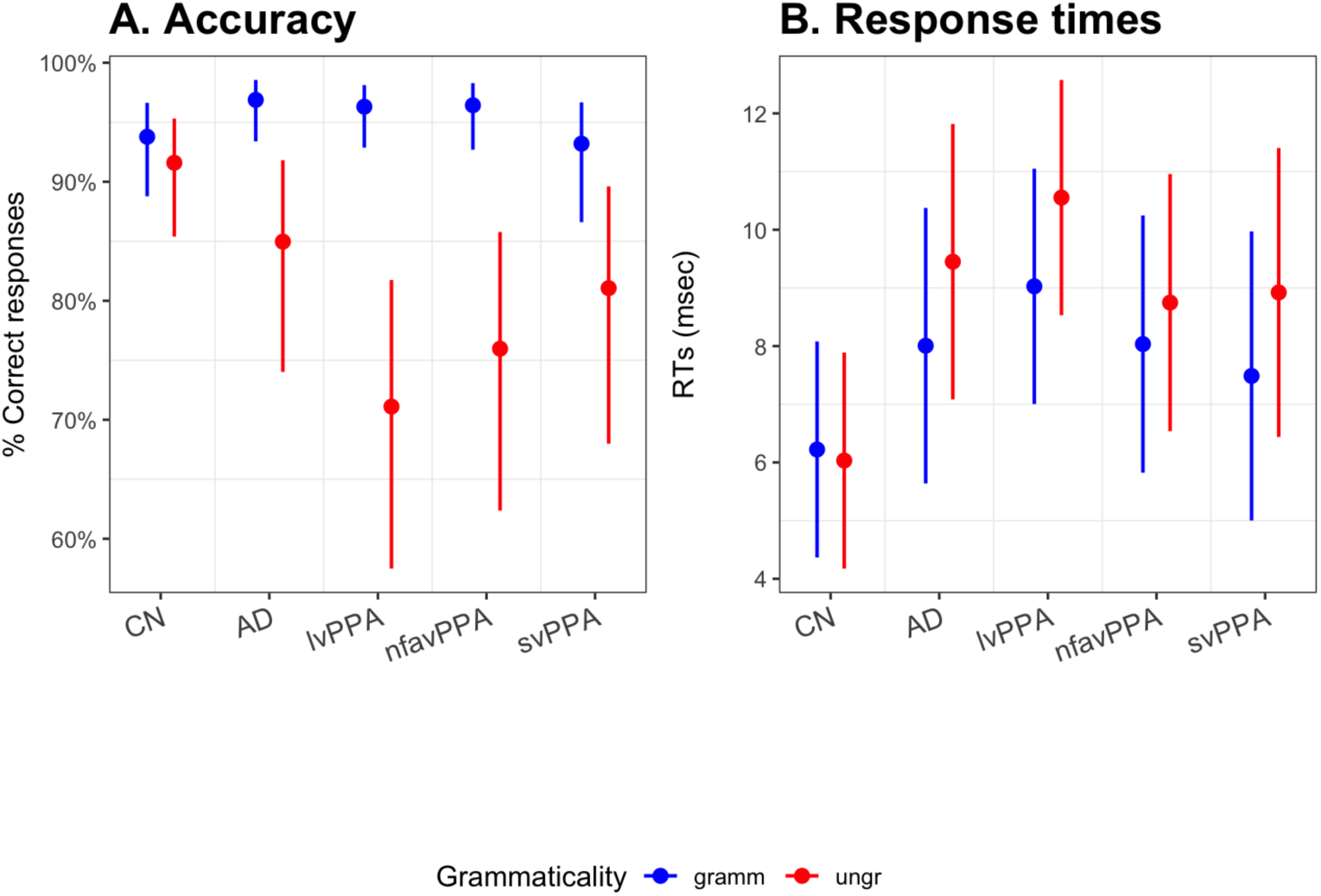
Predicted accuracy (panel A) and response times (panel B) across groups in grammatical and ungrammatical conditions. Dots indicate the point estimate; lines indicate standard error. Legend: CN: cognitively normal group; AD: Alzheimer’s Disease group; lvPPA: logopenic variant PPA group; nfvaPPA: non-fluent/agrammatic variant PPA group; svPPA: semantic variant PPA group.

#### Accuracy

LRTs showed that grammaticality and the interaction between group and grammaticality significantly increased model fit. Post-hoc analyses revealed that while the CN group was equally accurate in the evaluation of grammatical and ungrammatical sentences (grammatical: 0.94; ungrammatical: 0.92; odds ratio: 1.38; se: 0.506, z: 0.886, p= .3754), greater accuracy for correct compared to incorrect sentences was found in individuals with AD (grammatical: 0.97; ungrammatical: 0.85; odds ratio: 5.49; se: 2.188, z= 4.277, p <.001). Similar grammaticality effects emerged in the analysis of the three PPA variants (lvPPA: grammatical: 0.96; ungrammatical: 0.71; odds ratio: 10.59; se: 3.822, z = 6.536, p <.001; nfvaPPA: grammatical: 0.96; ungrammatical: 0.76; odds ratio: 8.53; se: 3.281, z = 5.575, p <.001; svPPA: grammatical: 0.93; ungrammatical: 0.81; odds ratio: 3.20; se: 1.191; z = 3.127; p= .0018). The inclusion of MMSE significantly increased model fit.

**Table 3.** Results of likelihood ratio tests for accuracy and response times. P-values are corrected using False Discovery Rate (Benjamini & Hochberg, 1995). Statistically significant results after correction are highlighted in bold.

| Accuracy | Df | Df diff | $\chi^2$ | Uncorrected p | Corrected p |
| --- | --- | --- | --- | --- | --- |
| Years of education+ years post onset | 5 |  |  |  |  |
| Group | 9 | 4 | 13.17 | .0105 | **.0140** |
| Grammaticality | 10 | 1 | 24.04 | <.000 | **<.000** |
| Group * Grammaticality | 14 | 4 | 42.71 | <.000 | **<.000** |
| MMSE | 15 | 1 | 5.61 | .0179 | **.0224** |
| Response times |  |  |  |  |  |
| Years of education+ age | 6 |  |  |  |  |
| Group | 10 | 4 | 12.59 | .0135 | **0.0269** |
| Grammaticality | 11 | 1 | 3.17 | .0752 | 0.1002 |
| Group * Grammaticality | 15 | 4 | 14.60 | .0056 | **0.0224** |
| MMSE | 16 | 1 | 1.57 | .2103 | 0.2103 |

*Legend. Df = degrees of freedom; Df diff. = difference in degrees of freedom*

The four diagnostic groups performed similarly to the CN group on grammatical sentences (*all ps* > .1) but differed in their performance on ungrammatical ones. Unlike AD, nfvaPPA and lvPPA were significantly less accurate than CN in evaluating incorrect sentences (AD vs. CN: z= 1.641, p= .2015; lvPPA vs. CN: z= 4.071, p= .0005; nfvaPPA vs. CN: z = 3.283, p = .005). Performance of svPPA in ungrammatical sentences marginally differed from CN (z = 2.296, p = .0654). Pairwise comparisons between clinical groups in both grammatical and ungrammatical trials did not reach statistical significance (all p*s* >.1).

#### Response times

Compared to the base model, grammaticality did not increase model fit, but group and its interaction with grammaticality did. Post-hoc analyses to disentangle the interaction revealed a grammaticality effect for the AD, lvPPA and svPPA groups, with longer latencies for responses to ungrammatical compared to grammatical trials (AD – grammatical: 8.01s; ungrammatical: 9.45 s; estimate: -1.444; se: 0.647, z=-2.233, p=0.0255; lvPPA – grammatical: 9.03 s; ungrammatical: 10.55s; estimate: - 1.525; se: 0.586, z=-2.603, p =.0092; svPPA – grammatical: 7.49s; ungrammatical: 8.92s; estimate: -1.435; se: 0.665, z=-2.157, p =.0310), but not for nfvaPPA (grammatical: 8.03s; ungrammatical: 8.75s; estimate: 0.713; se: 0.618, z: -1.155, p= .2482) and CN groups (grammatical: 6.22s; ungrammatical: 6.03 s; estimate: 0.190; se: 0.546, z: 0.347, p= .7284).

No significant differences were found among the five groups in grammatical trials (all p*s* > .05). In ungrammatical trials, longer latencies were found for lvPPA compared to the CN group (z=-3.206, p= .0135), while all other comparisons did not reach statistical significance (all p*s* >0.1). The inclusion of MMSE did not increase model fit.

### 4.3. Characterization of PPA variants and AD

#### Effect of cognitive functions on grammaticality judgment

##### Short-term memory

As illustrated in Table 4 and Figure 5, LRTs showed a three-way interaction between forward digits span, grammaticality and group driven by the different effect that cognitive scores had on the two levels of grammaticality in lvPPA and AD as opposed to nfvaPPA and svPPA. Post-hoc analyses revealed that forward digit span scores predicted the grammaticality effect significantly in lvPPA (estimate: -0.835, SE: 0.246, z: -3.389, p= .0007), marginally in the AD group (-estimate: 0.726, SE: 0.402, z: -1.806, p= .0709) but not in the nfvaPPA (estimate: -0.334, SE: 0.292, z: -1.144, p=.2525). The effect did not reach significance in the svPPA group either (estimate: 0.115, SE: 0.347, z: 0.332, p= .7399). In the lvPPA group, higher forward digit span scores predicted higher accuracy in ungrammatical trials.

**Figure 5.**
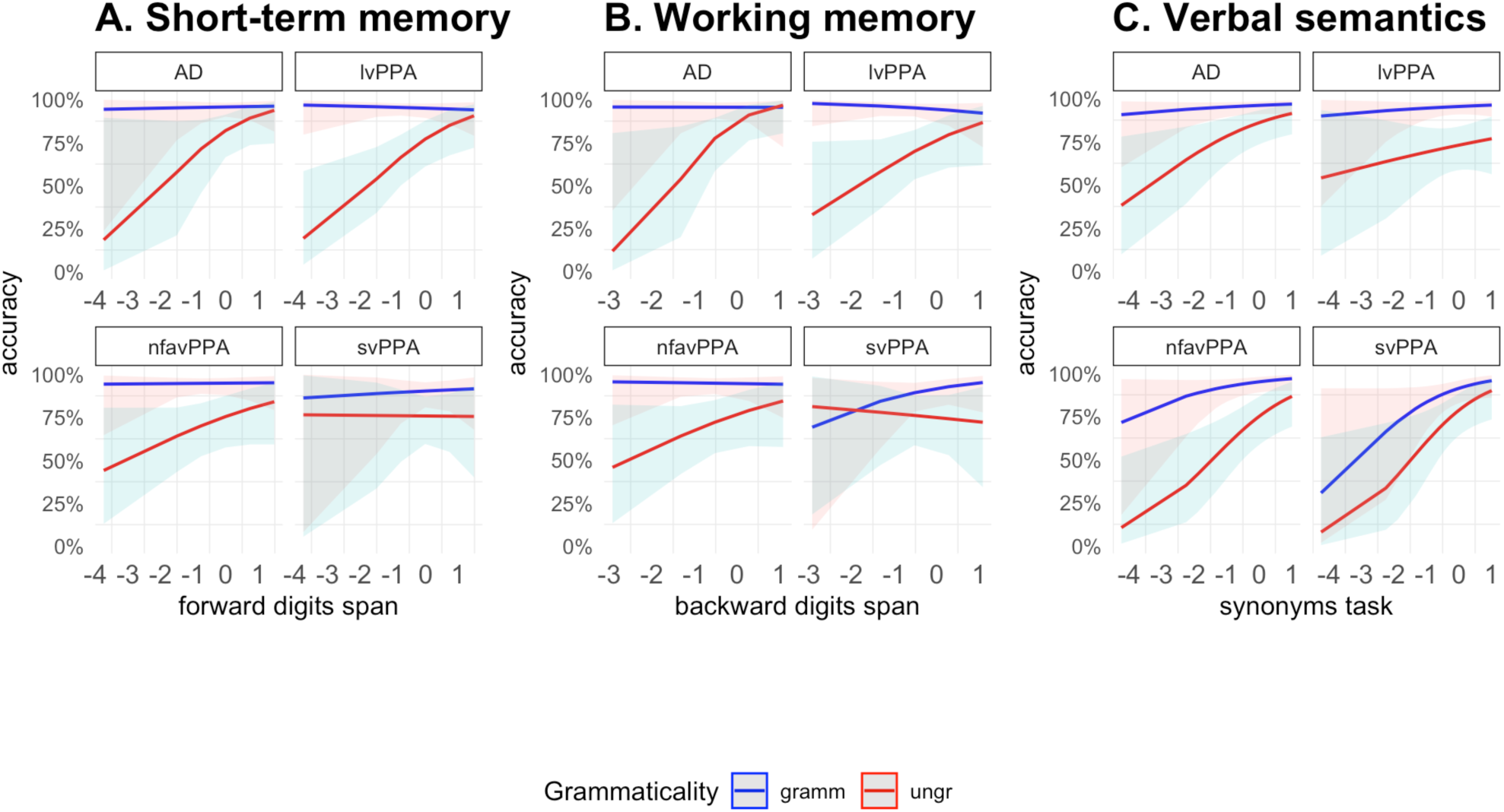
Predicted accuracy scores at different levels of performance on centered forward-digit span (panel A), backward digit span (panel B) and synonym task (panel C) for each diagnostic group. Legend: gramm = grammatical condition; ungr = ungrammatical condition.

Pairwise comparisons showed that the predictive effect of forward digit spans on grammaticality was similar across AD, lvPPA and nfvaPPA (all ps>.1), but it significantly differed between lvPPA and svPPA (estimate: -0.950, SE: 0.425, z: -2.233, p= .0255).

**Table 4.**
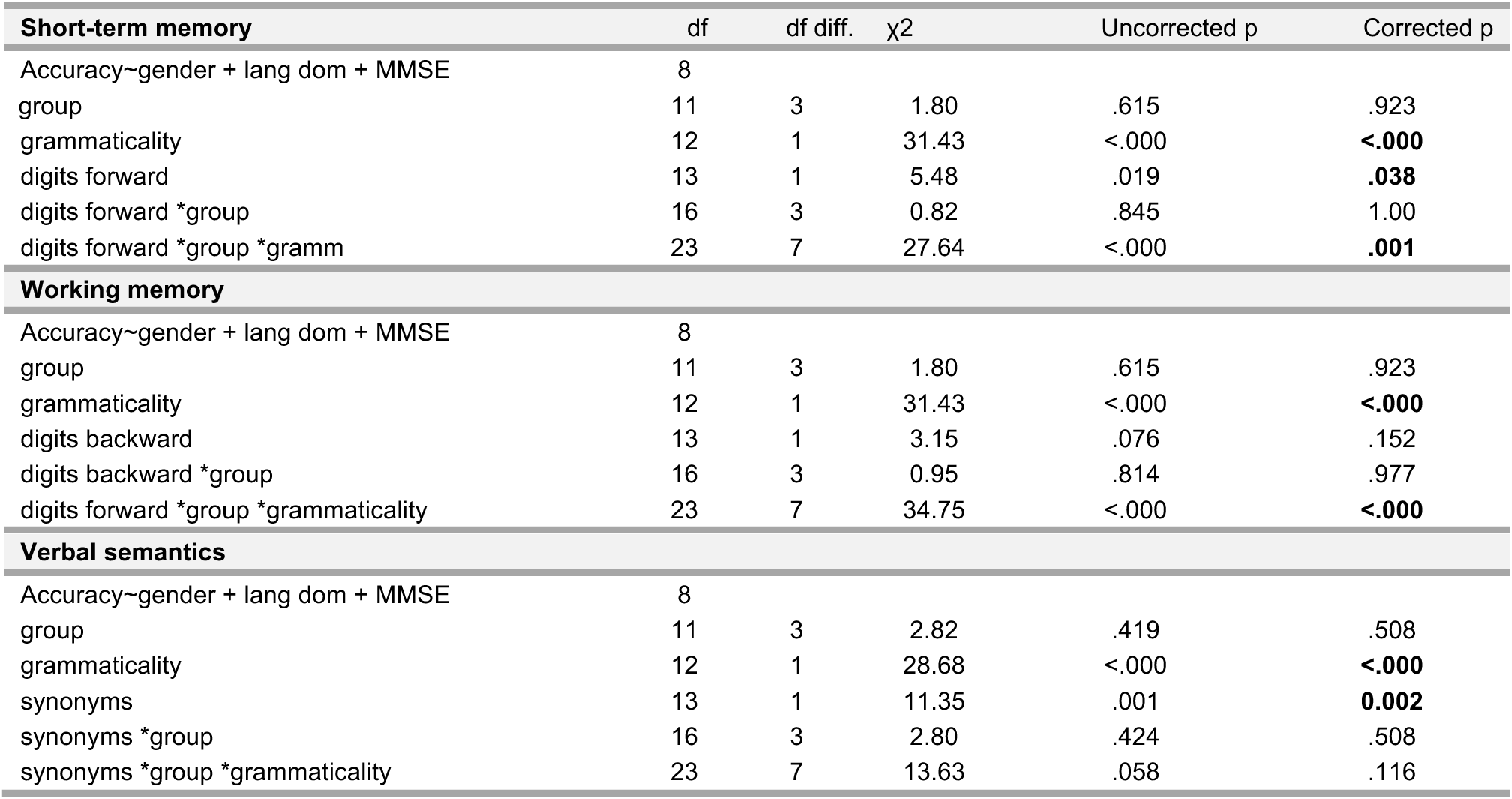
LRTs for the analysis of the effect of short-term memory, working memory and verbal semantics on grammaticality judgment accuracy. P-values are corrected using False Discovery Rate (Benjamini and Hochberg, 1995). Statistically significant results after correction are highlighted in bold.

#### Working memory

A three-way interaction between backward digit span, grammaticality and group emerged, driven by the different effect that this cognitive measure had on grammatical and ungrammatical conditions across groups. In the AD and lvPPA groups, the difference in accuracy between grammatical and ungrammatical conditions decreased as backward digit span scores improve (AD group - estimate: -1.366, SE: 0.515, z: -2.650, p=.0081; lvPPA - estimate: -0.977, SE: 0.302, z: -3.239, p=.0012). In both groups, higher digit span scores were associated with higher accuracy on ungrammatical conditions. Although not statistically significant, a similar trend emerged for the nfavPPA group (estimate: -0.538, SE: 0.374, z: -1.437, p= .1506). In the svPPA group, the difference in accuracy between grammatical and ungrammatical conditions increased with higher digit span scores (estimate: 0.675, SE: 0.338, z: 1.999, p=.0457).

Pairwise comparisons showed that the predictive effect of digit span on grammaticality was similar across AD, lvPPA and nfvaPPA (all p*s* > .2), but differed between svPPA and the other three clinical groups (AD - svPPA estimate: -2.041, SE: 0.616, z: -3.311, p=.0028; lvPPA – svPPA: estimate: - 1.652, SE: 0.453, z: -3.646, p=.0016; nfvaPPA – svPPA estimate: -1.213, SE: 0.504, z: -2.405, p=.0323).

#### Verbal semantics

Scores on the synonyms task had a similar positive effect on grammaticality judgement accuracy across groups, with accuracy increasing as performance on the synonyms task increased. This positive effect did not modulate performance on grammatical and ungrammatical conditions differentially.

### Accuracy across experimental conditions and groups

We report GLMEMs results for pairwise group comparison (AD vs. CN, lvPPA vs. CN, nfvaPPA vs. CN, svPPA vs. CN, see Figure 6).

**Figure 6.**
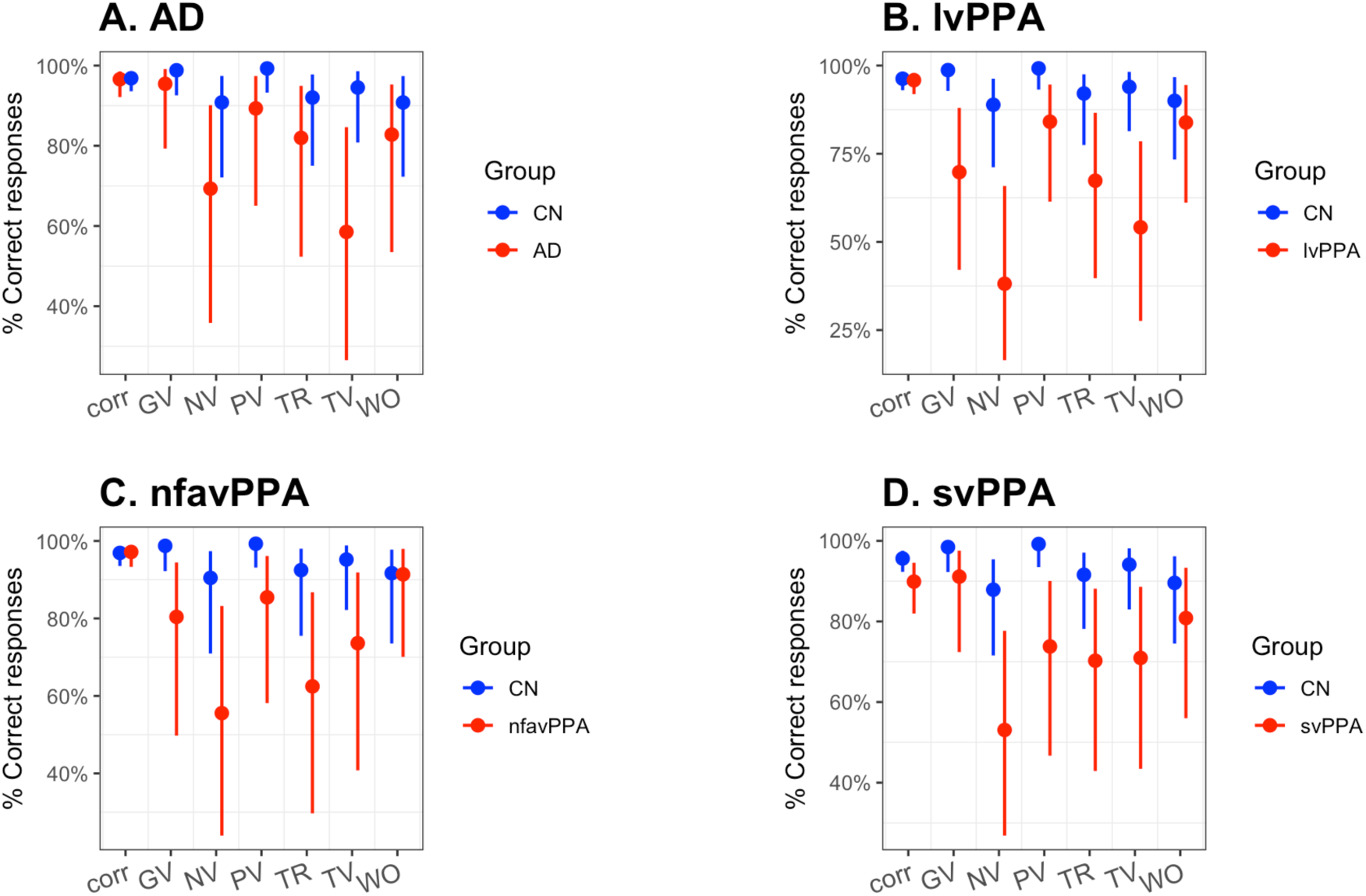
Predicted accuracy across groups and conditions. Legend: corr = correct; GV= gender violation; NV = number violation; PV = person violation; TV = tense violation; TR= transivity violation; WO = word order. AD: Alzheimer’s Disease; nfvaPPA: nonfluent/agrammatic variant PPA; lvPPA: logopenic variant PPA; svPPA: semantic variant PPA.

#### AD vs. CN

LRTs showed a significant effect of group (Chi-Square: 7.57, corrected p=.0089), condition (Chi-Square: 15.22, corrected p= .0186), and a significant interaction group x condition (Chi-Square: 21.81, corrected p=.0039). Post-hoc comparisons using estimated marginal means revealed that the CN group performed significantly better than the AD group in number agreement (estimate: 0.2283, SE: 0.1335, z: -2.527, p=.0115), person agreement (estimate: 0.0600, SE: 0.0691, z: -2.443, p=.0146) and tense agreement violations (estimate: 0.0817, SE: 0.0509, z: -4.018, p=.0001). The performance of the two groups did not differ in correct sentences, transitivity and word order violations (all p*s* >.1).

#### lvPPA vs. CN

LRTs showed a significant effect of group (Chi-Square: 20.72, corrected p= .00001), condition (Chi-Square: 25.95, corrected p= .00023), and a significant interaction group x condition (Chi-Square: 52.54, corrected p=.00000). Post-hoc comparisons revealed that the CN group outperformed the lvPPA group in the evaluation of gender (odds ratio: 33.15, SE: 28.049, z: 4.138, p<.0001), number (odds ratio: 12.96, SE: 6.626, z: 5.013, p<.0001), person (odds ratio: 23.91, SE: 26.181, z: 2.899, p=.0037), tense (odds ratio: 13.19, SE: 7.331, z: 4.641, p <.0001) and transitivity (odds ratio: 5.64, SE: 2.976, z: 3.277, p= .0011) but not in word order anomalies and correct sentences (all p*s* >.3).

#### nfvaPPA vs. CN

LRTs showed a significant group (Chi-square: 9.57, corrected p= .0030), condition (Chi-Square: 16.71, corrected p=.0104) and group x condition interaction (Chi-Square: 40.00, corrected p<.0001). Post-hoc comparisons to disentangle the interaction revealed that the CN group outperformed the nfvaPPA group in the following conditions: gender (odds ratio: 19.073, SE 16.949, z: 3.318, p= .0009), number (odds ratio: 7.596, SE 4.441, z: 3.469, p=.0005), person (odds ratio: 23.499, SE: 26.689, z: 2.780, p=.0054), tense (odds ratio: 7.116, SE: 4.596, z: 3.038, p=.0024) and transitivity anomalies (odds ratio: 7.382, SE: 4.417, z: 3.341, p=.0008). The two groups did not differ in the evaluation of correct and word-order anomalous sentences (all p*s* >.8).

#### svPPA vs. CN

LRTs revealed a significant group (Chi-Square: 16.48, corrected p=.0001), condition (Chi-Square: 13.87, corrected p= .0311) and group x condition interaction (Chi-Square: 15.42 p=.017197, corrected p= .0258). Post-hoc comparisons to disentangle the interaction showed that the CN group performed significantly better than the svPPA group in correct sentences (odds ratio: 2.45, SE: 0.906, z: 2.433, corrected p: .0150), number (odds ratio: 3.96, SE: 2.257, z: 2.418, p=.0156), person (odds ratio: odds ratio: 27.48, SE: 31.022, z: 2.935, p=.0033), tense (odds ratio: 4.02, SE: 2.584, z: 2.169, p=.0301) and transitivity anomalies (odds ratio: 4.61, SE: 2.625, z: 2.684, corrected p: .0073). No significant differences emerged between the two groups in word order conditions (p>.2).

### 4.4. Imaging correlates of experimental task performance

In the whole-brain VBM considering overall accuracy in the grammaticality judgement task (Figure 2, panel A), a large cluster centered in the left posterior temporal cortex was identified. The peak voxel was located in the left posterior middle temporal gyrus (*T* = 4.75), with additional local maxima in the left mid middle temporal gyrus (*T* = 4.75) and the left mid to posterior superior temporal gyrus (*T* = 4.59). Additional smaller clusters were observed in the left supramarginal gyrus (*T =* 4.46), left angular gyrus (*T =* 3.54), left planum temporale (*T =* 3.91), left superior frontal gyrus (*T =* 3.92), and right superior temporal gyrus (*T =* 3.76). The principal left temporal cluster located in the left middle temporal gyrus also survived cluster-level FWE correction (*p*FWE < .001).

Adding digit forward as an additional covariate in the VBM analysis produced a similar pattern (Figure 2, panel B), with a cluster located in the mid to posterior middle temporal gyrus, with three local maxima (*T* = 4.68, *T* = 4.63, *T* = 4.47). Smaller clusters were also identified in the left inferior temporal gyrus (*T* = 3.65), the more anterior part of the middle temporal gyrus (*T* = 3.52) and left supramarginal gyrus (*T* = 4.10). The left temporal cluster centered in the middle temporal gyrus also survived cluster-level FWE correction (*p*FWE = .001).

Similarly, adding digit backward as an additional covariate in the VBM analysis considering overall grammar accuracy (Figure 2, Panel C) led to left temporal cluster centered in the left middle temporal gyrus (peak *T* = 5.08). Additional local maxima in the left inferior temporal gyrus and adjacent white matter (*T* = 4.60) and the left posterior middle temporal gyrus (*T* = 4.59).

Additional local maxima within the cluster were observed within the left inferior temporal gyrus and underlying white matter (*T* = 4.60) and in the left posterior middle temporal gyrus (*T* = 4.59). Additional smaller clusters were observed in the left supramarginal gyrus (*T* = 4.07), left inferior temporal gyrus (*T* = 3.77), left cerebellum (*T* = 3.60), and right superior temporal gyrus (*T* = 3.47). The cluster centered in the left middle temporal gyrus also survived cluster-level FWE correction (*p*FWE < .001).

In the subgroup analysis restricted to participants with AD etiology (Figure 2, panel D), no suprathreshold clusters were identified. Suprathreshold peaks were observed in the left mid middle temporal gyrus (*T* = 4.52), in the left inferior temporal gyrus (*T* = 5.12), and the left cerebellum (*T* = 4.82).

In the subgroup of participants with FTLD etiology (Figure 2, panel E), whole-brain VBM analysis identified one cluster that survived family-wise error (FWE) correction at the cluster level (*p*FWE = .032). The peak voxel was located in the left mid temporal gyrus (*T* = 4.49), with additional local maxima in the left posterior middle temporal gyrus (*T* = 4.45) and left posterior superior temporal gyrus (*T* = 4.30). Suprathreshold peaks were observed in the left supramarginal gyrus (*T* = 4.88), in the left superior frontal gyrus (*T* = 4.76), in the left inferior frontal gyrus (*T* = 4.46) and the right inferior frontal gyrus (T = 4.28, T = 3.85). Complete statistical results for all VBM analyses thresholded at p < .001 (uncorrected, k ≥ 50 voxels), including peak MNI coordinates, cluster extent, T-statistics, and cluster- and voxel-level statistics, are reported in the Supplementary Materials 5.

## 5. Discussion

The goal of this study was to provide a comprehensive picture of morphosyntactic and transitivity comprehension in speakers of Spanish affected by PPA and AD. We designed a short, computer-based acceptability judgment task in which we manipulated the well-formedness of sentences to create inflectional, transitivity and within-constituent word order anomalies. We hypothesized that, if comprehension of these linguistic domains was impaired in both PPA and AD, the evaluation of sentence (un-)grammaticality should reliably elicit worse accuracy and longer response latencies compared to the control group.

Psychometric analyses revealed that our grammaticality judgment task could reliably detect the presence of impairment and that it has high internal consistency. Moreover, accuracy in the task moderately correlated with language and neuropsychological tests. Analysis of accuracy and RTs in grammatical and ungrammatical trials showed that, overall, both AD and PPA variants were found to be impaired in inflectional and transitivity comprehension, as revealed by the lower accuracy and longer latency with which participants evaluated sentence acceptability compared to the control group, while sensitivity to word order anomalies appeared more preserved. As expected, the four clinical groups retained the ability to correctly evaluate grammatical sentences but not to reject unacceptable ones. Analyses showed that short-term, working memory and verbal semantics functions were differentially associated with correct and incorrect sentence processing both within and across groups. While no statistically significant differences emerged among the four clinical groups, the lvPPA group showed the lowest accuracy scores, with difficulties especially evident in the analysis of number, gender and tense violations. VBM analyses indicated the left posterior temporal cortex as the main neuroanatomical correlate of grammaticality judgment performance across clinical groups. In the following, we discuss these findings and their implications in greater detail.

### 5.1. Psychometric properties

Correlations of accuracy with language and neuropsychological tests revealed moderate significant correlations, suggesting that the evaluation of sentence acceptability relies on language processing mechanisms that only partially overlap with those underlying sentence-picture matching and, sentence elicitation, and that recruit memory and cognitive mechanisms supporting processing speed only to a limited extent. In other words, evaluating morphosyntax and transitivity through a grammaticality judgment task valuably complements information obtained from other typical language tasks used in clinical and research practice without excessively taxing domain-general, non-linguistic functions.

A weak negative correlation emerged between response times and accuracy, suggesting that responses and their latencies do not consistently reflect a speed-accuracy tradeoff strategy, with higher latencies not always associated with higher accuracy and vice versa (Figure S1.1, Supplementary Material 1). In addition, response times did not reliably correlate with any linguistic or neuropsychological test. The lack of time pressure to provide a response, combined with the constant on-screen availability of the sentence, could have minimized the influence of memory and attention deficits in response latencies, thus resulting in non-significant correlations. Further studies with larger samples should corroborate this null finding and its interpretation.

Overall, ROC analyses revealed good discriminability (AUCs ranging from .74 to .85) across pairwise group comparisons, paired with moderate sensitivity, likely reflecting the wide range of morphosyntactic and transitivity impairment severity in our sample, spanning from severe impairment to performance within normal limits. This pattern was mostly pronounced in the lvPPA– CN group comparison, where sensitivity dropped to .62.

### 5.2. Grammatical-ungrammatical asymmetry and the nature of the impairment

In line with our predictions, all three PPA variants and the AD group showed impairment in the evaluation of sentence grammaticality, as evidenced by lower accuracy rates compared to the CN group. Deficits in the analysis of several types of sentential relations can be thus a common trait of these neurodegenerative conditions, in line with previous studies with PPA (Barbieri et al. 2021; Grossman et al. 2005; Lambon Ralph et al. 2012; Patterson et al. 2001; Price and Grossman, 2005; see Auclair-Ouellet, 2015 for a review) and AD patients (Fyndanis et al. 2013), including those testing comprehension with experimental paradigms other than offline grammaticality judgment, i.e. online word monitoring tasks. What this suggests is that PPA and AD overall morphosyntactic and transitivity impairment can be captured reliably also by offline analogic assessment tasks, thus reinforcing the usefulness of testing these linguistic abilities as part of routine language assessment together with other language tasks.

The data revealed robust asymmetry between accuracy in grammatical and ungrammatical trials in all clinical groups: both PPA and AD patients were accurate at detecting grammatical sentence well-formedness but performed worse than the CN group in ungrammatical conditions. This effect has been reported in previous studies with AD (Fyndanis et al. 2013;) and is also consistent with findings from post-stroke aphasia (Hagiwara, 1995; Varlokosta et al. 2006, Wilson & Saygin, 2004). A possible explanation for this asymmetric effect resides in the impairment of the cognitive functions involved in task performance, which could alter the type of processing heuristics adopted by patients to evaluate grammatical and ungrammatical sentences.

Psychometric analyses showed that overall accuracy on the grammaticality judgment task is positively associated with short-term and working memory, as measured by forward- and backward digit spans. This relation was further explored with regression models including the grammaticality factor, which revealed that, in AD and lvPPA, accuracy on ungrammatical trials increased as short-term and working memory functions improved. This is not surprising, as the detection of a mismatch involves checking the consistency between the current input, visible on screen, and an (internally generated) expected input. In ungrammatical conditions, reduced memory functions could determine unstable access and retrieval of expected inflectional and transitivity representations, leading to unstable conflict monitoring and “repair” mechanisms. This is in line with EEG findings by Barbieri et al. (2021), who showed late-onset and attenuated P600 effects for the lvPPA group, suggesting delayed sensitivity to, and arguably shallower repair of, the anomaly. Since lvPPA and AD show predominant parietal atrophy (Figure 1), this interpretation is also supported by neuroimaging studies with healthy participants that associated activity in parietal areas with conflict-monitoring operations involved in the evaluation of the match between the expected and the perceived stimulus (Botvinick, Cohen and Carter, 2004; Ye and Zhou, 2009), including when inflectional and case marking information are manipulated (Folia et al., 2009; Carreiras et al. 2015; Kuperberg et al., 2008, 2003; Mancini et al. 2017; Nieuwland et al., 2012; Quiñones et al. 2014, 2018).

The nfvaPPA group showed a trend towards a positive relation between accuracy on ungrammatical conditions and working memory impairment (Figure 5B), although it did not reach statistical significance. Memory impairments in nfvaPPA have been documented in association with inflectional impairment (Grossman et al. 2005; see Eikelboom et al. 2018 for a review of studies on working memory impairment in nfvaPPA) and related with fronto-insular patterns of atrophy, although the severity has been found to be comparably lower compared to lvPPA (Eikelboom et al. 2018). In line with these findings, the nfvaPPA group in this study showed a milder working memory impairment compared to the lvPPA group (Table 1), which arguably resulted in a weaker association between working memory and ungrammatical sentence accuracy.

A different scenario emerged in the svPPA group. A higher backward-digit span was associated with higher accuracy on grammatical sentences, suggesting that, in order to compensate for their semantic deficits, this group relies on working memory to evaluate sentences. Interestingly, this pattern does not occur in any of the other diagnostic groups, which showed at-ceiling performance on grammatical sentences. Although working memory impairment in svPPA has been reported in the literature (Eikelboom et al. 2018), future studies with a larger sample size should confirm this finding.

Verbal semantics was also found to be a significant predictor of accuracy. This cognitive function was evaluated by asking participants to select a word that best fitted the meaning of the target. Understanding the meaning of words involves accessing information contained in their lexico-syntactic representation, i.e., their syntactic category and inflectional paradigms, whether a verb is transitive, what arguments it takes. Better-preserved verbal semantic abilities may thus go hand in hand with stronger implicit lexico-syntactic knowledge, giving rise to a positive relationship between verbal semantics and accuracy that reflects an interconnection across multiple levels of linguistic analysis, namely the syntactic, inflectional, and argument selectional properties. This points to verbal semantics as a meaningful component of the cognitive processes underlying the parsing of both grammatical and ungrammatical sentences across groups.

Although only qualitatively (Table 4 and Figure 5), the effect of verbal semantics appears to differ across groups and levels of grammaticality. In the svPPA and nfvaPPA groups, accuracy in both grammatical and ungrammatical conditions showed a positive relation with verbal semantics, suggesting a more generalized impairment in the establishment of links across levels of analysis. In contrast, in the AD and lvPPA groups, verbal semantics was associated with accuracy in ungrammatical but not grammatical conditions. In these two groups, the ability to detect ungrammaticality thus appears to rely more heavily on preserved verbal semantics together with preserved short-term and working memory compared to other clinical groups.

Taken together, this set of analyses suggest that the performance of AD and the three PPA variants is comparable in terms of outcomes, i.e. they show a similar grammatical-ungrammatical asymmetry. However, the nature of the impairment underlying such an asymmetry could be different in AD-lvPPA vs. nfvaPPA-svPPA groups, as their different association with memory and verbal semantics suggest. However, given the small size of our clinical groups and the exploratory nature of these analyses, this set of results should be confirmed by larger-scale studies.

### 5.3. Accuracy across experimental conditions

Accuracy analysis across experimental conditions revealed no rigid boundary among clinical groups. In line with previous studies that reported impaired sensitivity to these features in English (Barbieri et al. 2021; Grossman et al. 2005; Kielar et al. 2018; Peele et al. 2007), the three PPA variants showed impaired sensitivity to tense and number agreement anomalies. Notably, accuracy on number anomalies was generally lower than on other ungrammatical conditions across PPA groups, falling below chance in the lvPPA group. This pattern may reflect the comparatively low visual and acoustic salience of number marking in Spanish, where ungrammaticality is signaled by the presence or absence of a single morpheme: -s in nouns and adjectives and -n in verbs (e.g., deportista**∅**SG vs. deportista***-sPL***, or corre-**∅SG** vs. corre***-nPL***). Such minimal cues stand in contrast to the more perceptually prominent differences that mark anomalies in other conditions e.g. person: el padre lleva/*lleva***ste***) and may therefore place disproportionate demands on the phonological and reading abilities known to be impaired in lvPPA (Brambati et al. 2009; Ramanan et al. 2022).

In line with our expectations, and in keeping with evidence from cognitively healthy speakers (Mancini et al. 2017; Quiñones et al. 2014, 2018), insensitivity to gender and person anomalies emerged in the semantic and logopenic variants of PPA. The different patterns of cortical atrophy that characterize these two variants suggest that poor accuracy in person and gender anomaly evaluation reflects the impairment of different underlying mechanisms. On the one hand, the difficulty in evaluating the ungrammaticality of these conditions in the lvPPA group could stem from core phonological (Wilson et al. 2014) and feature retrieval deficits (which also apply to number and tense violation impairment) derived from their short-term and working memory impairment. In contrast, patients diagnosed with svPPA could be impaired in the access to the semantic-conceptual information conveyed by nominal and verbal inflection. Interestingly, of the four clinical groups, only svPPA performed worse on correct sentences compared to the CN group. This is in line with our findings that different degrees of verbal semantics integrity is associated with processing of both grammatical and ungrammatical sentences in svPPA (Figure 5), as well as with fMRI studies in cognitively healthy Spanish speakers that showed involvement of the anterior temporal lobe during the processing of correct compared to anomalous sentences (Mancini et al. 2017; Quiñones et al. 2014).

Finally, the patterns of impairment evidenced in the AD group, person, tense and number agreement, closely align with previous studies in Greek (Fyndanis et al. 2013). The positive effect of working memory abilities on accuracy that we found suggests that AD’s inflectional comprehension impairment could stem from the interaction between their memory deficit and the progressive erosion of the semantic representation associated with inflectional information.

None of the clinical groups differed from CN participants in accuracy when evaluating word order anomalies, pointing to broadly preserved sensitivity to within-constituent phrase structure across all four groups. Yet, response times for this condition were significantly slowed in all groups relative to controls (Supplementary Materials 3), suggesting that while the outcome of the analysis process was preserved, the speed with which participants arrived at their judgments was not. This pattern of preserved accuracy alongside delayed responses may reflect effortful rather than automatic sensitivity to phrase structure well-formedness and it raises the possibility that outright insensitivity could emerge at more advanced stages of disease progression. Consistent with this interpretation, accuracy on word order anomalies appears numerically lower in patients with higher years post-onset (Supplementary Materials 4), though this observation remains exploratory, pending confirmation by a longitudinal study and/or a larger sample size.

### 5.4. Accuracy vs. response times (a-)symmetry

The analysis of response times revealed a scenario that only partially overlapped with the analysis of accuracy. A significant grammaticality effect emerged in AD, lvPPA and svPPA groups, with ungrammatical conditions eliciting longer latencies compared to grammatical ones. In these three groups, detecting ungrammaticality took longer and was more error-prone compared to evaluating grammatical stimuli, suggesting that participants were engaging with the task, in the attempt to find the source of the anomaly to repair it, but were struggling to do so efficiently. In contrast, in the nfvaPPA group, response latencies for grammatical and ungrammatical conditions were similar. In this group, the lower accuracy found on ungrammatical trials was not associated with longer response times for this condition compared to grammatical stimuli. A plausible interpretation for this finding is that individuals in the nfvaPPA group were not engaged in a deep analysis of the violation. This refrained them from finding the source of the anomaly and thus from repairing it, in line with the lack of P600 effect reported by Barbieri et al. (2021). In other words, while the other groups slowed down in the attempt to find the source of the uncertainty, the nfvaPPA group arguably lacked the cognitive control necessary to trigger the monitoring of response quality, due to frontal lobe atrophy. While theoretically plausible, more data from larger clinical groups are needed to corroborate these interpretations.

### 5.5. Imaging correlates of experimental task performance

Whole-brain VBM analyses consistently identified the left posterior temporal cortex as the principal neuroanatomical correlate of grammaticality judgment performance. Across all analyses, the strongest associations were centered on the posterior and middle portions of the left middle temporal gyrus, extending into the superior temporal gyrus.

This finding aligns with previous functional neuroimaging studies in healthy Spanish speakers showing recruitment of posterior temporal regions during the processing of morphosyntactic anomalies (Carreiras et al., 2015; Mancini et al., 2017; Quiñones et al., 2014, 2018), and with studies investigating the neural correlates of grammar processing in people with aphasia, showing that structural integrity of the left posterior temporal cortex predicts grammar performance both in primary progressive aphasia (e.g., Amici et al., 2007; Lorca-Puls et al., 2024) and in stroke aphasia (e.g., Dronkers et al., 2004; Matchin et al., 2022; Fahey et al., 2024; Biondo et al., 2024).

Importantly, the association between overall grammaticality judgement performance and posterior temporal regions remained significant after controlling for forward and backward digit span. Although our behavioral analyses demonstrated that short-term and working memory contribute to grammaticality judgment performance, particularly in AD and lvPPA, controlling for forward and backward digit span did not alter the principal anatomical correlates of grammar performance in the VBM analyses. This suggests that memory functions modulate successful performance but do not fully account for the relationship between posterior temporal atrophy and grammaticality judgment. Instead, posterior temporal integrity appears to contribute to grammatical processing beyond the variance explained by short-term auditory and working memory.

One potential concern is that the observed association reflects the temporoparietal atrophy characteristic of the AD spectrum, particularly lvPPA. To address this possibility, we repeated the analyses separately according to etiology. Importantly, the FTLD subgroup, composed of patients with nfvPPA and svPPA, also showed a significant cluster centered on the left posterior middle temporal gyrus. The persistence of posterior temporal correlates within the FTLD subgroup argues against the possibility that the whole-sample findings were driven solely by the inclusion of AD-spectrum patients and is consistent with recent VBM evidence demonstrating that receptive agrammatism in nfvPPA preferentially maps onto left posterior temporal cortex (Lorca-Puls et al., 2024), with a significant cluster centered on the left posterior middle temporal gyrus. This indicates that the association between grammaticality judgment performance and posterior temporal integrity is not solely attributable to the atrophy pattern of the AD-spectrum group, but extends across etiologies with markedly different patterns of neurodegeneration.

Despite previous functional imaging and lesion studies implicating left inferior frontal cortex in aspects of syntactic processing, no frontal regions emerged as independent structural correlates of grammaticality judgment performance in the present study. One possible explanation is that the grammaticality judgment task primarily required participants to detect morphosyntactic and verb-transitivity violations rather than resolve long-distance dependencies or construct highly complex hierarchical sentence representations. Under these task demands, successful performance may depend predominantly on posterior temporal regions supporting lexical-syntactic representations, whereas frontal mechanisms may become increasingly important when sentence comprehension requires manipulation of long-distance syntactic dependencies and greater working memory resources (Amici et al., 2007; Biondo et al., 2024; Lorca Puls et al., 2024; Matchin & Hickok, 2019; Rogalsky et al., 2008)

Finally, while our grammaticality judgment task differs from classic sentence to picture matching comprehension paradigms used in many previous studies, both tasks require analysis of morphosyntactic well-formedness during comprehension, suggesting that the posterior temporal cortex provides a common neural substrate for receptive grammatical processing across different task demands, as suggested by some neurocognitive language models (Matchin & Hickock, 2020). Together with our behavioral and psychometric data, VBM results are consistent with the view that overall grammaticality judgment performance reflects the interaction of language-specific representations and domain-general cognitive resources, rather than being reducible to memory impairment alone. More broadly, the present findings support the use of grammaticality judgment as a clinically informative tool for assessing receptive grammatical knowledge. Unlike sentence-picture matching tasks, whose performance may be influenced by visual processing, thematic role mapping, and executive demands associated with selecting among competing visual alternatives, grammaticality judgment isolates the evaluation of morphosyntactic well-formedness while preserving the need to process grammatical relations during comprehension. As such, it provides complementary information to conventional comprehension assessments and may be particularly useful for characterizing receptive grammatical impairment in neurodegenerative diseases.

### 5.6. Limitations

While this set of data have reliably shown that the analysis of inflectional and transitivity information can be impaired across the three PPA variants and AD, the relatively small size of the clinical and CN groups prevented us from characterizing the nature of the deficit and assessing the correlation between atrophy and accuracy patterns beyond purely hypothesis-generating findings. Future studies with bigger cohorts could adopt more specific statistical tests that enable the establishment of a clearer causal link between (in-)accurate morphosyntactic and transitivity analysis and specific cognitive functions (e.g., moderation analysis); (ii) the identification diagnosis-specific patterns in accuracy (e.g., with multinomial logistic regression and linear discriminant analysis); and (iii) the correlation between accuracy in experimental conditions and patterns of cortical atrophy.

Moreover, from a theoretical perspective, the offline nature of the task did not allow us to identify potential timing differences in the analysis of morphosyntactic and transitivity information. Paradigms that capture the time course underlying the reading and analysis process leading to the answer (e.g. eye tracking, eeg) could be used to cast light on this.

## 6. Conclusion

This study and its findings extend previous research on morphosyntactic and argument structure comprehension in PPA and AD, mostly English-oriented, to Spanish, a language whose richer inflectional system affords a finer-grained window into the nature and scope of grammatical impairment in these populations. By probing sensitivity to a wide spectrum of grammatical features, gender, number, tense, person, and transitivity, within a single, psychometrically validated paradigm, this study demonstrates that comprehension deficits in these domains are not a by-product of English morphological poverty but a robust feature of neurodegenerative language decline across languages. This has both theoretical and clinical implications. Theoretically, the cross-linguistic convergence of findings strengthens the case that morphosyntactic and transitivity impairment constitutes a shared dimension of language deterioration in PPA and AD, independent of the specific demands imposed by any one language. Clinically, it underscores the importance of developing assessment tools that are sensitive to the morphological properties of the language being tested, rather than adapting instruments designed for typologically different languages. The grammaticality judgment task introduced here offers a brief, reliable, and linguistically motivated means of detecting and characterizing comprehension impairment in Spanish-speaking populations, an understudied group in the neurodegenerative language literature. Future research should extend this approach to other morphologically rich languages and larger, longitudinal samples, to further clarify the trajectory of grammatical comprehension decline and its relationship to broader cognitive and neural deterioration across the PPA spectrum and in AD.

## Data Availability

All data produced in the present study are available upon reasonable request to the authors

## Acknowledgments

Simona Mancini acknowledges support from grants PID2024-159519OB-I00 from the Spanish State Research Agency, CEX2020-001010/AEI/10.13039/501100011033 (Severo Ochoa Excellence) and BERC 2022-2025 Program to BCBL. Miguel Santos-Santos acknowledges support from the Spanish Institute of Health Carlos III co-funded by the European Union (Juan Rodés research grant JR18-00018; Fondo de investigación sanitaria grant PI19/00882), the Alzheimer’s Association clinician scientist fellowship (AACSF-22-972945), and the National Institutes of Health (R01AG080470). The SPIN cohort received funding from the Fondo de Investigaciones Sanitario (FIS), Instituto de Salud Carlos III (PI13/01532, PI14/01126, PI16/01825, PI17/01019, PI17/01896, PI18/00335, PI18/00435, PI19/00882, PI20/01473, PI20/00836,PI21/00791,PI21/01395, PI21/00063, PI21/00791, PI22/00611, PI22/00307, PI24/00598, PI24/00968, PI24/01087, INT19/00016, INT23/00048, AC19/00103, DTS22/00111, PI25/00422) and the CIBERNED program (Program 1, Alzheimer Disease to AL), jointly funded by Fondo Europeo de Desarrollo Regional, Unión Europea, “Una manera de hacer Europa”. The SPIN cohort was also supported by the National Institutes of Health (NIA grants 1R01AG056850-01A1; R21AG056974; R01AG080470; and R01AG061566), by Generalitat de Catalunya (2017-SGR-547, SLT006/17/125, SLT006/17/119, SLT002/16/408, SLT042/25/000034), “Marató TV3” foundation grants 20141210, 044412 and 20142610, a grant from the Fundació Bancaria La Caixa to RB (DABNI project), Fundació Catalana Síndrome de Down and Fundació Víctor Grífols i Lucas. Horizon 21 Consortium is partly funded by Jérôme Lejeune Foundation. We acknowledge the Support for Research Groups funding from the Department of Research and Universities from the Generalitat de Catalunya (2021 SGR 00979). Stephanie Grasso acknowledges support from the National Institute on Aging of the National Institutes of Health (R01AG080470) and the Alzheimer’s Association (24AARG-D-1246996).

## Author contribution

**Simona Mancini**: Conceptualization, formal analysis, methodology, software, visualization, writing (original draft, review and editing). **Nicoletta Biondo**: formal analysis, software, writing (original draft, review and editing). **Marco Calabria**: conceptualization, resources, writing (review and editing). **Clara Martin**: conceptualization, writing (review and editing). **Sonia Marqués-Kiderle**: data curation, project administration, resources, writing (review and editing). **Estefanía García Hernandez**: data curation, project administration, resources, writing (review and editing). **Júlia Filella Mercé**: data curation, project administration, resources, writing (review and editing). **Camille Wagner Rodriguez**: data curation, project administration, resources, writing (review and editing). **Judit Selma González**: data curation, project administration, resources, writing (review and editing). **Jesús García Castro**: data curation, project administration, resources, writing (review and editing). **Sara Rubio Guerra**: data curation, project administration, resources, writing (review and editing). **Isabel Sala**: data curation, project administration, resources, writing (review and editing). **María Belén Sánchez Sandinós**: data curation, project administration, resources, writing (review and editing). **Ignacio Illán-Gala**: data curation, funding acquisition, project administration, resources, writing (review and editing). **Alexandre Bejanin**: data curation, project administration, resources, writing (review and editing). **Alberto Lleó**: data curation, funding acquisition, project administration, resources, writing (review and editing). **Juan Fortea**: data curation, funding acquisition, project administration, resources, writing (review and editing). **Stephanie Grasso**: funding acquisition, writing (review and editing). **Miguel Santos Santos**: conceptualization, data curation, funding acquisition, investigation, methodology, project administration, resources, supervision, writing (original draft, review and editing).

## Footnotes

1 Unless otherwise specified, we use ‘morphosyntax’ to refer to both constituent structure and inflection.

2 Across studies, patients could also differ in their underlying pathology. Some patients diagnosed with svPPA have underlying Pick Disease, associated with more fronto-parietal atrophy, instead of TDP pathology (Spinelli et al. 2017)

