## Supplemental Material for "A Spanish grammaticality judgment task for neurodegenerative diseases: inflectional, transitivity, and word order comprehension in PPA and Alzheimer’s Disease"

### Supplementary Materials

#### Contents


Supplementary Material 1: Correlation between grammaticality judgment and response times 1

Supplementary Materials 2: Effect of sentence length on accuracy 2

Supplementary Materials 3: Condition effect in response times 4

Supplementary Materials 4: Effect of years post onset on accuracy 7

Supplementary Materials 5: Output of the VBM analyses 8

#### Supplementary Material 1: Correlation between grammaticality judgment and response times

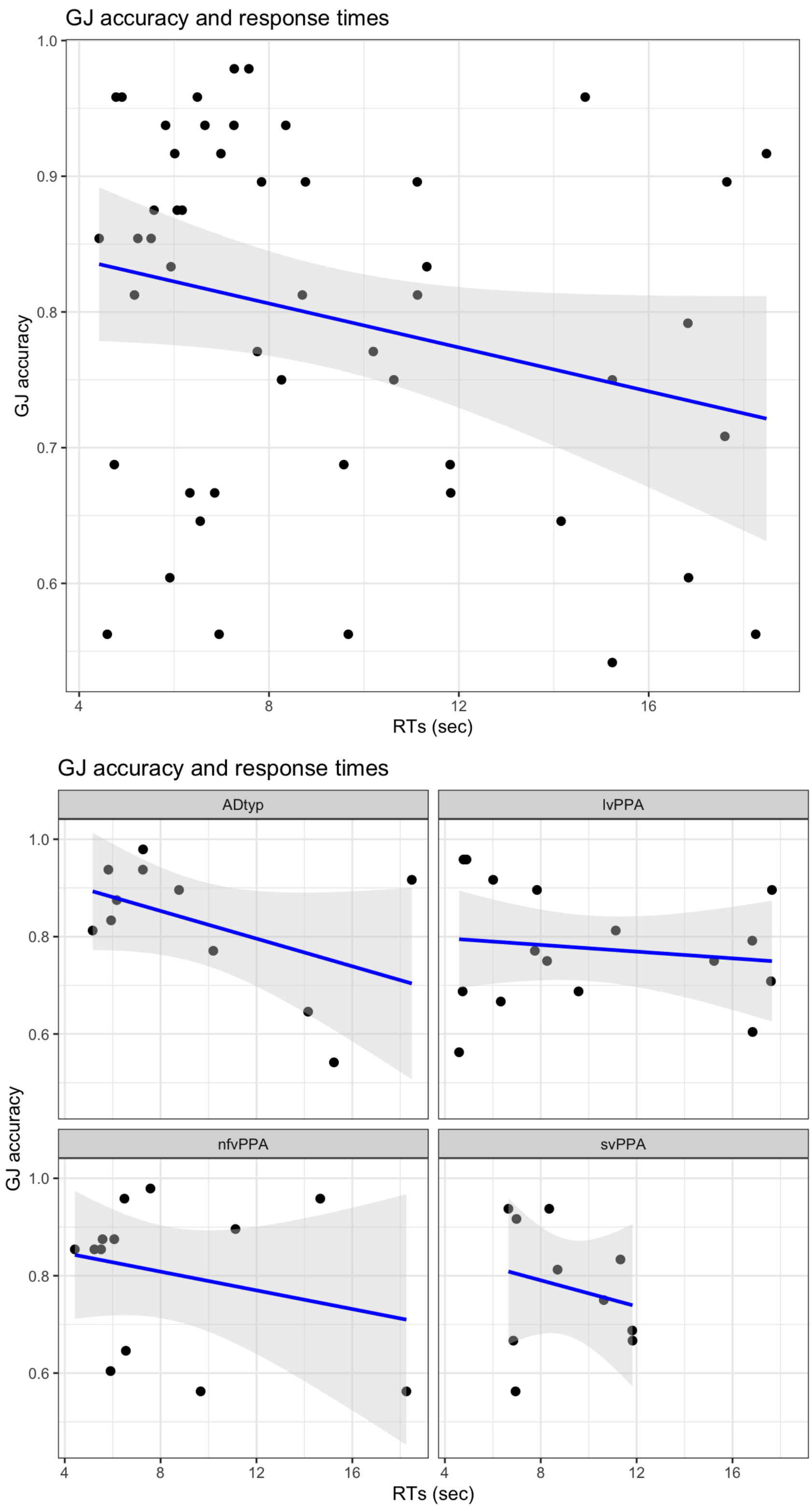


*Figure S1.1. Correlation between grammaticality judgment (GJ) accuracy and response times (RTs)*

#### Supplementary Materials 2: Effect of sentence length on accuracy

*Data analysis*

Three glmer models including sentence length (measured in number of words per sentence) and its interaction with group and grammaticality (length* group and length*group*grammaticality) were sequentially added to the series of models used for the main analysis of accuracy and response times (see Section 4.2). Random effect structure included by-subject and by-item intercepts. Results from likelihood ratio tests (LRTs) were FDR adjusted and reported below. Results from estimated marginal means are illustrated in Figure S3.1.

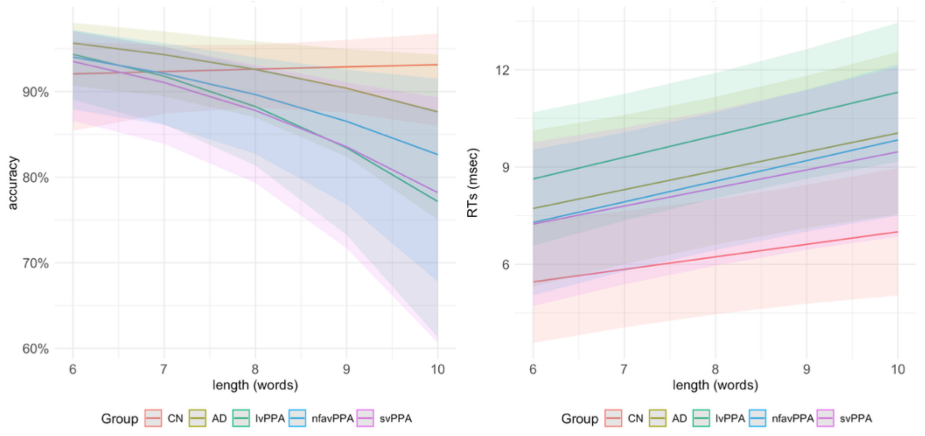


***Figure S2.1***. *Effect of sentence length (in words) on accuracy (left panel) and RTs (right panel) across groups*

*Results*

Accuracy. The inclusion of sentence length ($\chi^2$ =5.74, adjusted p=.0209), as well as its interaction with group ($\chi^2$ = 14.32, adjusted p=.0148) increased model fit, but not the three-way interaction with grammaticality ($\chi^2$ = 3.74, adjusted p=.5873). Pairwise post-hoc comparisons revealed that the effect of sentence length in the four clinical groups differed from that of the control group (CN vs. AD: estimate: .3230, SE: 0.135, z: 2.386, p=.0425; CN vs. lvPPA: estimate: 0.4390, SE: 0.122, z: 3.600, p=.0032; CN vs. nfvaPPA: estimate: 0.3388, SE: 0.129, z: 2.618, p=.0294; CN vs. svPPA: estimate: 0.3873, SE: .131, z: 2.965, p=.0151). The effect of sentence length on accuracy did not differ among clinical groups (all ps>.6).

RTs. The inclusion of sentence length improved model fit ($\chi^2 = 11.21$, adjusted p= .0057), such that response times increased as sentence length increased. However, the effect of this predictor was independent from group ($\chi^2 = 2.46$, adjusted p=.7615) and grammaticality ($\chi^2 = 2.29$, adjusted p=.8071).

#### Supplementary Materials 3: Condition effect in response times

*Methods*

The effect of each experimental condition on response times was modeled using linear mixed effect models with Condition (7 levels: correct, gender, number, person, tense, verb-argument structure and word order violations) as fixed factor, along with group and their interaction. The simplest model included age and years of education. Random effect structure included by-subject and by-item intercepts.

*Results*

*AD vs. CN*. LRTs showed a significant effect of group (Chi-Square: 12.92, p= .000326, adjusted p=.0010) but not of condition (Chi-Square: 1.45, p= .962529, adjusted p= .9625). A significant interaction group x condition emerged (Chi-Square:17.93, p=.006406, adjusted p=.0096). Post-hoc comparisons using estimated marginal means revealed that the CN group had significantly faster response times compared to the AD group in all conditions: correct sentences (estimate: -3.36, SE: 1.11, t: -3.018, p=.0047), gender (estimate: -3.33, SE: 1.41, t: -2.362, p=.0204), number (estimate: -6.00, SE: 1.41, t: -4.250, p=.0001), person (estimate: -5.60, SE: 1.41 z: -3.969, p=.0001), tense (estimate: -4.73, SE: 1.41, t: -3.355, p=.0012), verb-argument structure (estimate: -4.73, SE: 1.41, t: -3.355, p=.0012) and word order (estimate: -3.78, SE: 1.41, t: -2.678, p=.0088).

*lvPPA vs. CN*. LRTs showed a significant effect of group (Chi-Square: 14.01, p=.000181, adjusted p=.00054) but not of condition (Chi-Square: 4.28, p=.639116, adjusted p=.63912). The interaction between group and condition significantly increased model fit (Chi-Square: 14.50 p=.024529, adjusted p=.03679). Post-hoc comparisons revealed that the CN group were significantly faster than the lvPPA group in the evaluation of all conditions: correct sentences (estimate: -3.83, SE: 1.19, z: -3.209, p=.0026), gender (estimate: -5.00; SE: 1.44, t: -3.466, p=.0008), number (estimate: -6.34, SE: 1.44, t: -4.395, p<.0001), person (estimate: -5.99, SE: 1.44, t: -4.151, p=.0001), tense (estimate: -4.89, SE: 1.44, t: -3.389, p= .0011), verb-argument structure (estimate: -4.82, SE: 1.44, t: -3.337, p= .0013) and word order (estimate: -6.24, SE: 1.44, t: -4.327, p<.0001).

*nfvaPPA vs. CN*. LRTs showed a significant group (Chi-square: 7.98, p=.004736, adjusted p= .0142) but not of condition (Chi-Square: 3.88, p= .692547, adjusted p=0.6925). The interaction between group and condition significantly increased model fit (Chi-Square: 15.65, p=.015740, adjusted p=.0236). Post-hoc comparisons to disentangle the interaction revealed that the CN group was faster than the nfvaPPA group across all conditions: correct sentences: (estimate: -2.47 SE: 1.01, t: -2.434, p=.0202), gender (estimate: -3.13, SE: 1.15, t: -2.724, p=.0085), number (estimate: -2.52, SE: 1.15, t:-2.194, p=.0323), person (estimate: -3.32, SE: 1.15, t:-2.890, p=.0054), tense (estimate: -4.68, SE: 1.15, t: -4.079, p=.0001), verb-argument structure anomalies (estimate: -2.80, SE: 1.15, -2.434, p=.0181) and word order (estimate: -3.78, SE: 1.15, t:-3.291, p=.0017) anomalies.

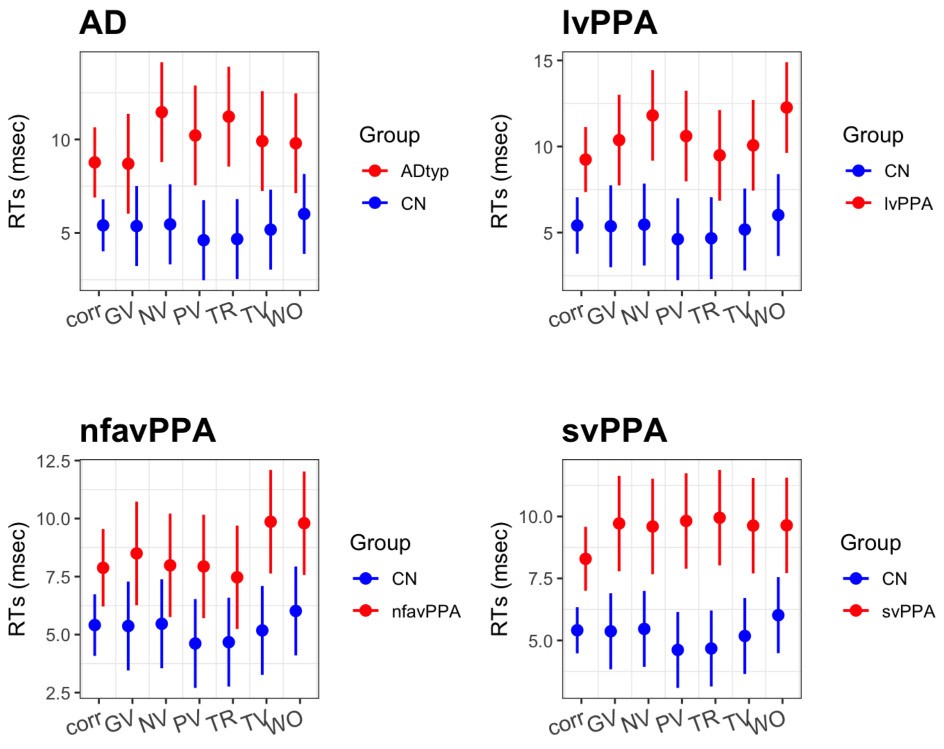


***Figure S3.1*** Predicted response times across conditions and groups.

svPPA vs. CN. LRTs revealed a significant effect of group (Chi-Square: 20.17 p=.000007, adjusted p<.001) but not of condition (Chi-Square: 1.85, p= .933101, adjusted p=.9331). The interaction between group and condition increase model fit (Chi-Square: 21.23, p=.001666, adjusted p=.0025). Post-hoc comparisons to disentangle the interaction showed that the CN group was faster than the svPPA group across all conditions: correct sentences: (estimate: -2.88 , SE: 0.743, t: -3.877, p=.0004), gender (estimate: -4.35, SE: 0.971, t: -4.481, p= <.0001), number (estimate: -4.13, SE: 0.971, t: -4.258, p<.0001), person (estimate: -5.20, SE: 0.971, t:-5.362, p<.0001), tense (estimate: -4.45, SE: 0.971, t: -4.588, p<.0001), verb-argument structure anomalies (estimate: -5.28, SE: 0.971, t:-5.437 p<.0001) and word order (estimate: -3.62, SE: 0.971, t:-3.734, p=.0003) anomalies.

#### Supplementary Materials 4: effect of years post onset on accuracy

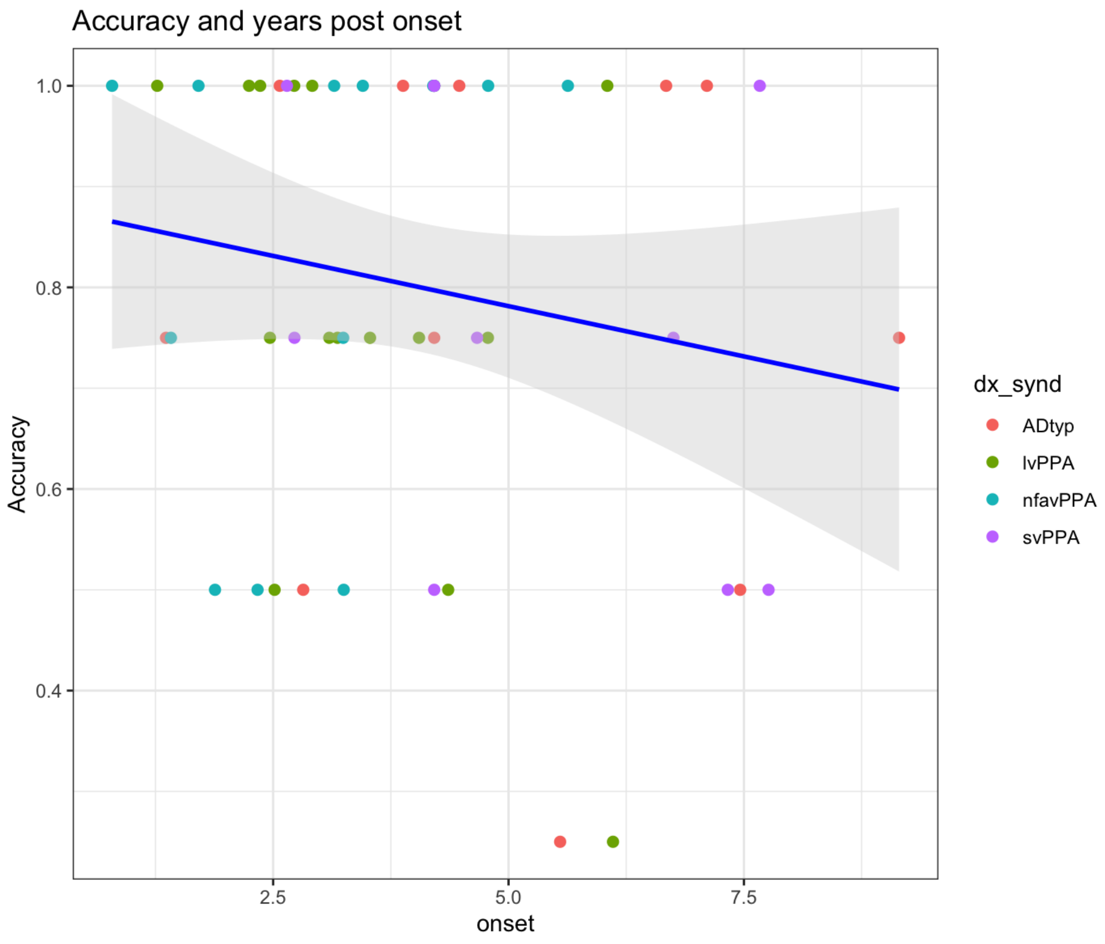


***Figure S4.2***. Accuracy in the word-order anomaly condition as a  function of years post onset.

#### Supplementary Materials 5: output of the VBM analyses

The tables below report complete statistical results for all VBM analyses thresholded at $p < .001$ (uncorrected, $k \geq 50$ voxels), including peak MNI coordinates, cluster extent, $T$-statistics, and cluster- and voxel-level statistics.

***Table S5A (Overall grammaticality judgment accuracy)***

| *Cluster* | *Cluster pFWE* | *k* | *Cluster puncorr* | *Peak pFWE* | *T* | *Peak puncorr* | *MNI coordinates [x, y, z]* |
|---|---|---|---|---|---|---|---|
| **1** | <.001 | 7233 | <.001 | .080 | 4.75 | <.001 | [-64 -42 2] |
| | | | | .080 | 4.75 | <.001 | [-58 -21 -9] |
| | | | | .121 | 4.59 | <.001 | [-57 -10 -6] |
| **2** | .423 | 329 | .179 | .168 | 4.46 | <.001 | [-50 -28 42] |
| **3** | .710 | 126 | .404 | .515 | 3.92 | <.001 | [-16 52 32] |
| | | | | .812 | 3.57 | <.001 | [-27 57 27] |
| **4** | .815 | 67 | .552 | .523 | 3.91 | <.001 | [-50 -39 18] |
| **5** | .655 | 159 | .347 | .657 | 3.76 | <.001 | [63 -14 -6] |
| **6** | .784 | 84 | .501 | .829 | 3.54 | <.001 | [-46 -70 42] |

***Table S5B (Overall accuracy controlling for digit forward)***

| *Cluster* | *Cluster pFWE* | *k* | *Cluster puncorr* | *Peak pFWE* | *T* | *Peak puncorr* | *MNI coordinates [x, y, z]* |
|---|---|---|---|---|---|---|---|
| **1** | .001 | 3470 | <.001 | .108 | 4.68 | <.001 | [-69 -32 -4] |
| | | | | .121 | 4.63 | <.001 | [-66 -40 0] |

| | | | | | | | |
|---|---|---|---|---|---|---|---|
| | | | | .177 | 4.47 | <.001 | [-57 -42 -2] |
| **2** | .800 | 78 | .509 | .396 | 4.10 | <.001 | [-50 -27 40] |
| **3** | .714 | 125 | .396 | .770 | 3.65 | <.001 | [-39 -6 -39] |
| | | | | .863 | 3.52 | .001 | [-46 -4 -36] |
| | | | | .927 | 3.39 | .001 | [-46 0 -46] |
| **4** | .744 | 108 | .432 | .862 | 3.52 | .001 | [-58 0 -24] |

***Table S5C (Overall accuracy controlling for digit backward)***

| *Cluster* | *Cluster pFWE* | *k* | *Cluster puncorr* | *Peak pFWE* | *T* | *Peak puncorr* | *MNI coordinates [x, y, z]* |
|---|---|---|---|---|---|---|---|
| **1** | <.001 | 3899 | <.001 | .037 | 5.08 | <.001 | [-69 -39 0] |
| | | | | .129 | 4.60 | <.001 | [-44 -34 -18] |
| | | | | .129 | 4.59 | <.001 | [-57 -44 -2] |
| **2** | .796 | 78 | .516 | .407 | 4.07 | <.001 | [-50 -28 40] |
| **3** | .541 | 234 | .252 | .664 | 3.77 | <.001 | [-39 -6 -39] |
| | | | | .687 | 3.74 | <.001 | [-46 -4 -36] |
| **4** | .618 | 182 | .312 | .797 | 3.60 | <.001 | [-36 -74 -34] |
| **5** | .835 | 57 | .584 | .880 | 3.47 | .001 | [54 -12 -10] |

***Table S5D (Overall accuracy in AD etiology group)***

| *Cluster* | *Cluster pFWE* | *k* | *Cluster puncorr* | *Peak pFWE* | *T* | *Peak puncorr* | *MNI coordinates [x, y, z]* |
|---|---|---|---|---|---|---|---|

| | | | | | | | |
|---|---|---|---|---|---|---|---|
| **1** | .629 | 175 | .282 | .177 | 5.12 | <.001 | [-46 -33 -21] |
| **2** | .495 | 258 | .194 | .287 | 4.82 | <.001 | [-39 -84 -22] |
| **3** | .356 | 368 | .125 | .452 | 4.52 | <.001 | [-58 -48 2] |
| | | | | .635 | 4.23 | <.001 | [-66 -45 4] |

***Table S5E (Overall accuracy in FTLD etiology group)***

| *Cluster* | *Cluster pFWE* | *k* | *Cluster puncorr* | *Peak pFWE* | *T* | *Peak puncorr* | *MNI coordinates [x, y, z]* |
|---|---|---|---|---|---|---|---|
| **1** | .705 | 135 | .308 | .353 | 4.88 | <.001 | [-50 -28 42] |
| | | | | .745 | 4.22 | <.001 | [-58 -30 46] |
| **2** | .646 | 164 | .262 | .414 | 4.76 | <.001 | [-27 51 32] |
| | | | | .942 | 3.78 | .001 | [-16 51 33] |
| **3** | .167 | 570 | .046 | .499 | 4.61 | <.001 | [-50 -56 12] |
| | | | | .652 | 4.37 | <.001 | [-57 -46 48] |
| | | | | .726 | 4.25 | <.001 | [-56 -56 22] |
| **4** | .032 | 1116 | .008 | .574 | 4.49 | <.001 | [-58 -26 -10] |
| | | | | .602 | 4.45 | <.001 | [-66 -39 0] |
| | | | | .693 | 4.30 | <.001 | [-56 -32 3] |
| **5** | .730 | 123 | .331 | .596 | 4.46 | <.001 | [-44 22 -10] |
| **6** | .831 | 76 | .448 | .602 | 4.45 | <.001 | [-66 -28 32] |
| **7** | .745 | 116 | .345 | .705 | 4.28 | <.001 | [40 26 8] |

| | | | | | | | |
|---|---|---|---|---|---|---|---|
| | | | | .829 | 4.07 | <.001 | [51 22 9] |
| **8** | .885 | 50 | .545 | .920 | 3.85 | .001 | [44 46 -9] |
| | | | | .945 | 3.77 | .001 | [50 44 -14] |
